# Genomic ascertainment to quantify prevalence and cancer risk in adults with pathogenic and likely pathogenic germline variants in RASopathy genes

**DOI:** 10.1101/2024.10.09.24314324

**Authors:** Jung Kim, Gina Ney, Megan N. Frone, Jeremy S. Haley, Uyenlinh L. Mirshahi, Esteban Astiazaran-Symonds, Mariya Shandrina, Gretchen Urban, H. Shanker Rao, Rick Stahl, Alicia Golden, Marielle E. Yohe, Andrea M. Gross, Yi Ding, David J. Carey, Bruce D. Gelb, Douglas R. Stewart

## Abstract

**Purpose:** Genomic ascertainment of electronic health record-linked exome data in two large biobanks was used to quantify germline pathogenic/likely pathogenic (P/LP) variant prevalence, cancer prevalence, and survival in adults with non-*NF1* RAS/mitogen-activated protein kinase genes (RASopathies).

**Patients and Methods:** Germline RASopathy variants were examined from adult participants in UK Biobank (UKBB; n=469,802), Geisinger MyCode (n=167,050) and Mount Sinai Bio*Me* (n=30,470). Variants were classified as per American College of Medical Genetics/Association for Molecular Pathology criteria and reviewed by a RASopathy variant expert. Heterozygotes harbored a RASopathy pathogenic/likely pathogenic variant; controls harbored wild type or benign/likely benign RASopathy variation. To distinguish germline variants from clonal hematopoiesis, benign tissues were Sanger sequenced. Tumor phenotype and demographic data were retrieved from MyCode and UKBB.

**Results:** Pathogenic variants in Noonan syndrome-associated genes (excluding known Noonan syndrome with multiple lentigines variants) were the most common with an estimated prevalence that ranged between 1:1,772–1:3,330 in the three cohorts. Pathogenic variants in cardiofaciocutaneous syndrome-associated genes had an estimated prevalence of 1:41,762– 1:55,683 in two cohorts. Pathogenic variants in *SPRED1* (Legius syndrome) were more frequent in UKBB (1:19,567 [95%CI: 1:13,150–1:29,116]) compared to MyCode (1:41,762 [95%CI: 1:15,185–1:130,367]). In *SPRED1-*heterozygotes, cancer prevalence was significantly increased in UKBB (OR:3.8 [95% CI: 2.48–8.64]; p=1.2×10^-3^) but not in the MyCode cohort. Pathogenic variants in *HRAS* (Costello syndrome) were not identified. In MyCode and UKBB cohorts, there was no significant increase in cancer prevalence in individuals with Noonan-, *CBL-* and CFC syndrome-associated pathogenic variants.

**Conclusion:** Genomic ascertainment from two large biobanks did not show evidence of elevated cancer risk in adult Noonan syndrome heterozygotes. There may be an increased cancer risk for adult *SPRED1* heterozygotes.

## Introduction

The RASopathies are a set of disorders that arise from germline pathogenic variation in genes within the RAS/mitogen-activated protein kinase (MAPK) pathway, leading to hyperactivation of downstream effectors. In turn, the dysregulation of this pathways affects many aspects of cellular function including growth, proliferation, and apoptosis, causing a range of clinically recognizable phenotypes. There are clinical similarities across the RASopathies, including neurocognitive/developmental delay, craniofacial anomalies, skin manifestations, cardiac disease and short stature^1^. Given the rarity of several of the RASopathies and variable expressivity, it is consequently difficult to estimate prevalence^2^. Thus, a genome-first approach to characterize and estimate prevalence of these syndromes is a logical step to improve detection of patients with mild or novel phenotypes and help better define disease penetrance. Moreover, some common features associated with RASopathies, such as short stature, can be captured from the electronic health record (EHR) and used to describe associated clinical phenotypes^3^. This strategy allows the correlation of relevant genotypes with phenotypic data from EHRs, without bias from results of those individuals with a clinical diagnosis.

Previously, Wenger *et al*. analyzed pathogenic variants in 12 genes associated with the RASopathies in the Mount Sinai’s BioMe Biobank (Bio*Me*) cohort (n = 32,344) and the UK Biobank (UKBB) (n = 49,960)^4^. From both cohorts, a total of 21 individuals harbored a pathogenic (P) or likely pathogenic (LP) RASopathy variant. Of those, just three (14%) individuals had been diagnosed with a RASopathy, yet half of the individuals harboring a pathogenic variant demonstrated ≥ 1 classic Noonan syndrome (NS) feature. Well-described NS features, such as short stature and cardiac anomalies, were less frequent than expected, however other conditions, such as hypothyroidism and autoimmune disorders, were significantly enriched compared to controls^4^. This and similar analyses highlight how utilizing a genome-first strategy can refine penetrance estimates and uncover unexpected clinical phenotypes in rare disorders.

Hyperactivating somatic variants in RAS/MAPK genes are among the most frequent oncogenic mutations in human cancer and represent molecular therapeutic targets. Cancer risk in the RASopathies is well-documented in certain disorders (especially in childhood and adolescence)^5–7^ based on phenotypic and familial ascertainment. It is known that cancer risk is elevated in Costello syndrome, where there is high cancer incidence, early onset of bladder cancer, and elevated risk for pediatric neuroblastoma and rhabdomyosarcoma^5,8^. Notably, elevated cancer risk also exists in specific high-risk NS variants, which are associated with increased risk for juvenile myelomonocytic leukemia, neuroblastoma, and rhabdomyosarcoma^5^. In other disorders, however, such as cardiofaciocutaneous (CFC) and Legius syndrome, cancer risk is unknown^9^. Moreover, despite case reports of cancer in adults with germline P/LP variants in RAS/MAPK genes^10^, cancer prevalence and outcomes in adults with germline RAS/MAPK variation is unknown. In this study, we applied a genome-first approach to interrogate the exome sequence of individuals in three large population and health system-based cohorts to quantify germline P/LP variant prevalence and examined cancer prevalence and survival of adults with non-*NF1* RAS/MAPK P/LP variants.

## Methods

### Cohorts

The UKBB is a large population-based prospective study of participants aged 40–69 years at recruitment, with extensive matching phenotypic and genomic data including 469,802 participants with exome data^11^. The DiscovEHR study consists of a subset of individuals who consented to participate in the Geisinger MyCode Community Health Initiative^12^ who had exome sequence and EHR-linked data. Exome sequencing was performed in collaboration with the Regeneron Genetics Center on a total of 170,503 participants. Detailed descriptions of MyCode study design and collection of phenotypic and genotypic data have been published previously^12^. A subset of 170,503 who are ≥ 18 years old was selected for this study (n = 167,050). This study was approved by the Geisinger Institutional Review Board. The Bio*Me* Biobank is an ancestrally diverse, EHR-linked biobank of over 55,000 patients enrolled from ambulatory care practices across the Mount Sinai Health System in New York City. The present study population consisted of 30,470 Bio*Me* participants (of whom 30,129 are ≥ 18 years old) with available research exome sequence data were used in this study^13^.

### Genes and syndromes

RASopathy genes were selected based on ClinGen RASopathy Gene Curation Expert Panel (GCEP) gene-disease associations^14^. Genes with classifications that were disputed as of 2/13/23 were excluded from analyses. The selected non-*NF1* RASopathy genes were then clustered into five recognized syndromes based on known RASopathy associations: Casitas B-lineage lymphoma *(CBL*) syndrome, CFC syndrome, Costello syndrome, NS, Noonan syndrome with multiple lentigines (NSML) and Legius syndrome (**Supplementary Table 1**).

### Variant filtering and annotation

From all three biobanks, variants with a genotype quality ≤ 30, read depth ≤ 10X, ABHet ≤ 0.25, ABHet ≥ 0.75, or alternate allele read ≤ 3 were removed. Variants were annotated with snpEFF^15^, ClinVar (database retrieved 12/08/2022) and InterVar (v.2.1.2)^16^ as previously described^17^. As most P/LP RASopathy gene variants are gain-of-function, variants predicted as loss-of-function (*e.g.,* stop-gained, frameshift, splice donor/acceptor) were excluded, except for *SPRED1*, for which loss-of-function is a known disease-causing mechanism. As a final step, all loss-of-function variants in *SPRED1* were reviewed on Integrative Genome Viewer.

### Variant classification

Variants were classified in a hierarchical manner, where ClinVar classifications took priority followed by InterVar, as previously described^18^. For the analysis, we selected variants that were P/LP by ClinVar or InterVar. As a final step, all P/LP variants were reviewed by B.D.G., a member of ClinGen’s RASopathy GCEP and Variant Classification Expert Panel (VCEP), according to consensus guidelines for variant interpretation. Variants determined to be P/LP after expert review were included in the analysis (**Supplementary Table 2**). Individuals without P/LP variants classified by InterVar/ClinVar in any of 20 RASopathy genes of interest were classified as controls.

### Sanger sequencing, clonal hematopoiesis and selection of variants

It can be difficult to distinguish true germline variants in RASopathy genes from clonal hematopoiesis (CH) in blood-derived exome sequences^19,20^. We Sanger sequenced DNA from 25 benign tissues from individuals in the MyCode cohort who had variants that are known to appear at high frequencies (> 5%) in CH (*CBL, KRAS, NRAS, PTPN11*) or are less likely to occur in CH (*SOS1, BRAF*) (**Supplementary Table 3).** Germline variants (variants detected in both blood and a sequenced benign tissue) were confirmed in individuals < 60 years of age and variant allele frequency (VAF) > 0.4 (**Supplementary Figure 1**). Conversely, increasing age at blood draw (> 60 years) and lower variant allele fraction (VAF < 0.4) correlated with an increased likelihood that a variant was of somatic origin (**Supplementary Figure 1**). Since it was not feasible to sequence samples for all variants or sequence any samples from UKBB and Bio*Me*, all variants in *CBL, KRAS, NRAS,* and *PTPN11* were excluded in any participant where VAF was ≤ 0.4 and age at sample collection was ≥ 60 years (n excluded = 32 (UKBB); 34 (MyCode); 6 (Bio*Me*)). To avoid the potential inclusion of CH variants in the other 16 RASopathy genes, “stringent” filtering (VAF ≤ 0.4 and age at sample collection ≥ 60) excluded all variants in all genes with these criteria. **Supplementary Figure 2** summarizes the approach to minimize the effects of CH in this study. **Supplementary Table 2** lists all P/LP variants included in the analysis.

### Phenotypic data

Cancer phenotypes were extracted from each of the biobanks for all cases and controls. For UKBB, fields 41270/41271 (Diagnoses – ICD10/9), 40001 (Underlying cause of death), and 40006/40013 (Type of cancer: ICD10/9) were extracted on 10/16/2023. For MyCode, the EHR and the Geisinger Cancer Registry were queried; for Bio*Me*, the Mount Sinai Data Warehouse was queried to obtain ICD10 codes. For cancer phenotype analysis, any diagnoses made at less than 18 years of age were eliminated from the analysis. For each biobank, cancer cases were identified via extraction of all ICD10 C codes and ICD9 140-208 codes (including all cancer diagnoses). Furthermore, to exclude potential somatic variants circulating in blood, anyone with ICD10 codes C42 (hematopoietic and reticuloendothelial system), C81-96/200-208 (malignant neoplasm, stated or presumed to be primary, of lymphoid, hematopoietic and related tissue), D46 (myelodysplastic syndrome) and D47 (other neoplasms of uncertain behavior of lymphoid, hematopoietic and related tissue) were excluded. NS diagnosis was queried using ICD10 code Q87 (“Other specified congenital malformation syndromes affecting multiple systems”), which may be used to code non-NF1 RASopathies. Height z-scores were calculated as previously described for UKBB^4^. For Geisinger, the median height of all height measurements taken after 18 years were used to calculate the height z-score.

### Statistical analyses

Statistical analysis were performed using R version 4.1.0 and SAS Enterprise Guide v8.3.0.103. Demographic comparison was performed using t-test for age, body mass index (BMI), and height, and Fisher exact test for sex, smoking history, cancer, death, and Q87 code. Cancer risk odds ratio was modeled using logistic regression with age, sex, smoking history, and BMI as covariates. Overall survival and time-to-cancer were plotted using the survival and ggplot2 packages and cox-proportional hazard model was modeled with age, sex, smoking history, self-reported race, and BMI as covariates. Restricted mean age was calculated using the rmean function. Lollipop plots were generated using trackViewer; oncoprint was generated using ComplexHeatmap packages. Power estimates were performed as previous described in Chow et al.^21^

### Power to detect predisposition to common and rare cancers in UK Biobank, MyCode and BioMe

**Supplementary** Figures 3-5 shows power as a function of presumed true odds ratio for a range of cancer rates in the UKBB (**Supplementary Figure 3**), MyCode (**Supplementary Figure 4**) and Bio*Me* (**Supplementary Figure 5**) using cohort-specific prevalences from **Table 1**.

**Table 1.** Estimated prevalence of germline RASopathy Pathogenic/Likely Pathogenic variants in three biobanks. Stringent: excludes all variants in genes with ABHet ≤0.4 and age at blood draw ≥60 years. CI: 95% confidence interval.

|  |  | UK Biobank<br>(469,618 exomes) |  | Geisinger MyCode<br>(167,050 exomes) |  | Mount Sinai BioMe<br>(30,129 exomes) |  |
| --- | --- | --- | --- | --- | --- | --- | --- |
|  |  | count | frequency | count | frequency | count | freq |
| *CBL* |  | 14 | 1:33,544 (1:19,982–<br>1:56,309) | 8 | 1:20,881 (1:10,581–<br>1:41,207) | 0 | - |
| CFC | stringent | 9 | 1:52,179 (1:27,453–<br>1:99,178) | 3 | 1:55,683 (1:17,474–<br>1:215,605) | 0 | - |
|  |  | 10 | 1:46,961 (1:25,509–<br>1:86,453) | 4 | 1:41,762 (1:15,185–<br>1:130,367) | 0 | - |
| Noonan | stringent | 141 | 1:3,330 (1:2,824–<br>1:3,927) | 68 | 1:2,456 (1:1,938–<br>1:3,113) | 15 | 1:2,008 (1:1,217–<br>1:3,314) |
|  |  | 149 | 1:3,151 (1:2,684–<br>1:3,700) | 73 | 1:2,288 (1:1,820–<br>1:2,876) | 17 | 1:1,772 (1:1,106–<br>1:2,838) |
| NSML |  | 21 | 1:22,362 (1:14,627–<br>1:34,188) | 7 | 1:23,864 (1:11,560–<br>1:49,264) | 1 | 1:30,129 (1:4,639–<br>1:577,181) |
| Noonan<br>without<br>NSML | stringent | 120 | 1:3,913 (1:3,273–<br>1:4,679) | 61 | 1:2,738 (1:2,132–<br>1:3,517) | 14 | 1:2,152 (1:1,282–<br>1:3,612) |
|  |  | 128 | 1:3,668 (1:3,086–<br>1:4,361) | 66 | 1:2,531 (1:1,989–<br>1:3,219) | 16 | 1:1,883 (1:1,159–<br>1:3,058) |
| Legius | stringent | 24 | 1:19,567 (1:13,150–<br>1:29,116) | 3 | 1:55,683 (1:17,474–<br>1:215,605) | 0 | - |
|  |  | 24 | 1:19,567 (1:13,150–<br>1:29,116) | 4 | 1:41,762 (1:15,185–<br>1:130,367) | 0 | - |
| Costello |  | 0 | - | 0 | - | 0 | - |

Regardless of the stringency, for NS, there was over 90% power to detect all (22% rate) and common (12%) cancers with an odds ratio of > 2 in UKBB. In MyCode, there was 79% power and 66% power to detect this excess risk in all and common cancers, respectively. There was very limited power to detect excess risk in NS in Bio*Me*. In UKBB and Geisinger, power to observe excess risk in the rarest cancers (< 1%) and in CFC and *SPRED1* was very limited.

### A note on terminology

It is important to draw a distinction between a person who harbors (*i.e.*, is heterozygous for) a RASopathy P/LP variant versus an individual who has been diagnosed with a RASopathy syndrome (*i.e.,* harbors a RASopathy P/LP variant and is penetrant); with EHR data, it is not always feasible to distinguish these two possibilities. Individuals in the first category will be referred to as “heterozygotes” or “RASopathy heterozygotes” (see **Supplementary Table 1** for gene- and syndrome-specific terms), and those in the second will be referred to, when appropriate, as having a RASopathy syndrome. The analysis of *PTPN11* heterozygotes harboring variants known to be associated with NSML was limited to descriptive statistics (prevalence, oncoprint) given the limited number of observed events (cancer and death).

## Results

### Prevalence of germline P/LP variants in RASopathy genes in adults

The prevalence of germline P/LP variants in genes associated with each syndrome (**Supplementary Table 1**) was quantified in the three biobanks (**Table 1**). Variants in NS-associated genes were the most common with an estimated prevalence of 1:1,772 to 1:3,330, consistent with previously published estimates^22^. Since specific *PTPN11* variants in our cohort are associated with NSML, we also estimated its prevalence (1:22,362–1:30,129) and NS-associated genes without known NSML-associated variants (1:1,883–1:3,913). Of the genes investigated in the Bio*Me* cohort, only P/LP variants in NS genes were detected. About 45% of the P/LP variants in NS genes were in *PTPN11*, consistent with previous findings^23^ (**Supplementary Figure 6**A). Pathogenic variants in genes associated with CFC syndrome, a rare RASopathy with a typically more severe clinical presentation^2^, were more frequent in UKBB and MyCode (1:41,762 to 1:55,683; **Table 1**). Variants in *BRAF*, which account for approximately 50% of CFC diagnoses, made up half (n = 4/10 in UKBB; 2/4 in MyCode) of the variants associated with CFC in this analysis **(Supplementary Figure 6**B). The difference in P/LP variant frequency in *SPRED1* (Legius syndrome) in the UKBB (1:19,567) and MyCode cohorts (1:41,762) (**Table 1**)^24^ is not explained by read depth differences in *SPRED1* in the two cohorts (MyCode: 31.2; UK Biobank: 17.7). *HRAS* P/LP variants (Costello syndrome) were not identified in any of the three biobanks.

### No difference in demographic measures between heterozygotes and controls

**Supplementary Table 4** lists demographic (covariate) data and p-values for differences between CFC, *CBL*, NS, NS without NSML, NSML and *SPRED1* heterozygotes vs. controls for MyCode and UKBB. Generally, there were very minimal differences in age, sex, smoking and BMI between heterozygotes and controls.

### Stature and Q87 code frequency in RASopathy heterozygotes vs. controls

Since short stature is a known feature of some RASopathies, height z-score was calculated from available biobank data. Across all syndromes, a wide range (-3.4 to 2.8) of height z-scores was observed (**Supplementary Figure 7**). No significant differences in height z-scores were observed in CFC or *CBL* heterozygotes vs. controls in the UKBB or MyCode cohorts (**Supplementary Table 4)**. However, significantly lower height z-scores were observed in NS- and NS-without-NSML heterozygotes (UKBB and MyCode), NSML heterozygotes (UKBB) and *SPRED1* heterozygotes (UKBB) compared to controls (**Supplementary Table 4)** but no data on growth hormone use was available. Furthermore, NSML heterozygotes had lower height z-scores compared to controls but were taller, on average, than NS heterozygotes. No RASopathy heterozygote in the MyCode and Bio*Me* cohorts carried a ICD10 Q87 code; six (4%) NS- heterozygotes (none of them NSML heterozygotes) in UKBB had a Q87 code.

### Cancer risk in adult RASopathy heterozygotes

Cancer prevalence in adults in MyCode and UKBB was calculated as an odds ratio in RASopathy heterozygotes vs. controls. Demographic data were not available for controls in Bio*Me* to assess cancer incidence for this biobank. In both MyCode and UKBB, there was no significant increase in cancer prevalence in NS heterozygotes vs. controls (**Figure 1)**. This was also true for *CBL* and CFC heterozygotes (**Figure 1**), although the number of cancer-case observations was smaller. In UKBB, *SPRED1* heterozygotes had a significantly increased cancer prevalence compared to controls (OR: 3.8; 95% CI: 2.48–8.64; p = 1.2×10^-3^; **Figure 1**). In MyCode, there was a small number of observations (n = 4) with a non-significant difference (p = 0.98) in cancer prevalence in *SPRED1*-heterozygotes.

**Figure 1.**
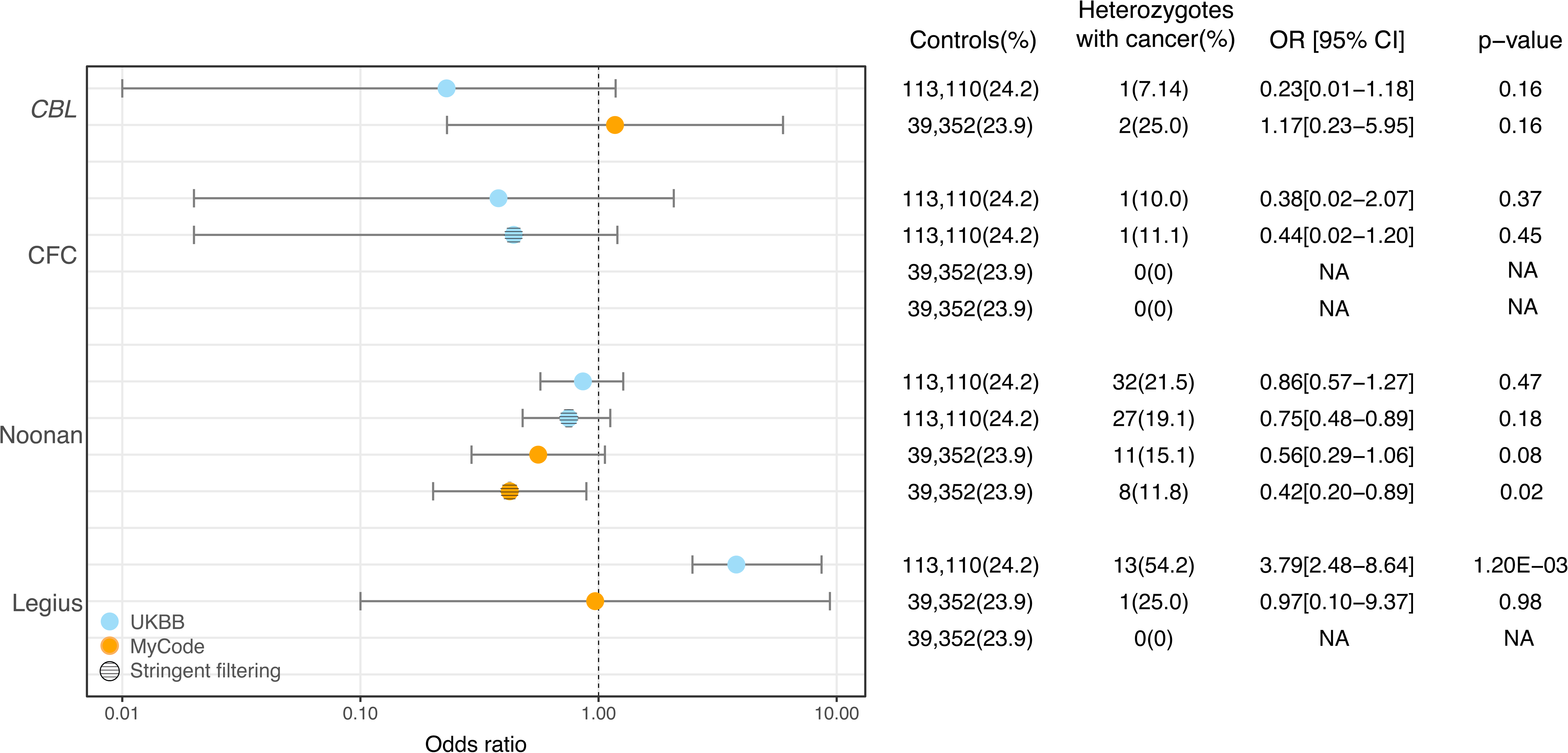
Cancer risk in *CBL-*, CFC-, Noonan- and *SPRED1*-heterozygotes in MyCode and UKBB cohorts with and without stringent filtering.

Figure 2 shows cancer type in RASopathy heterozygotes from the three cohorts. The most common cancer observed across all (except *PTPN11* heterozygotes) of the gene-syndrome groupings was malignancy of the skin. In *PTPN11* heterozygotes, gastrointestinal malignancies were the most common. Of the four NSML heterozygotes with cancer, all had malignant melanoma (age at diagnosis range from 41-73) and two of four cases had lentigo maligna histology. Notably, the cancer types included many common adult-onset solid tumors, including breast, lung, and gastrointestinal cancers. Gene-specific maps depicting variants and cancer are shown in **Supplementary Figure 8**.

**Figure 2.**
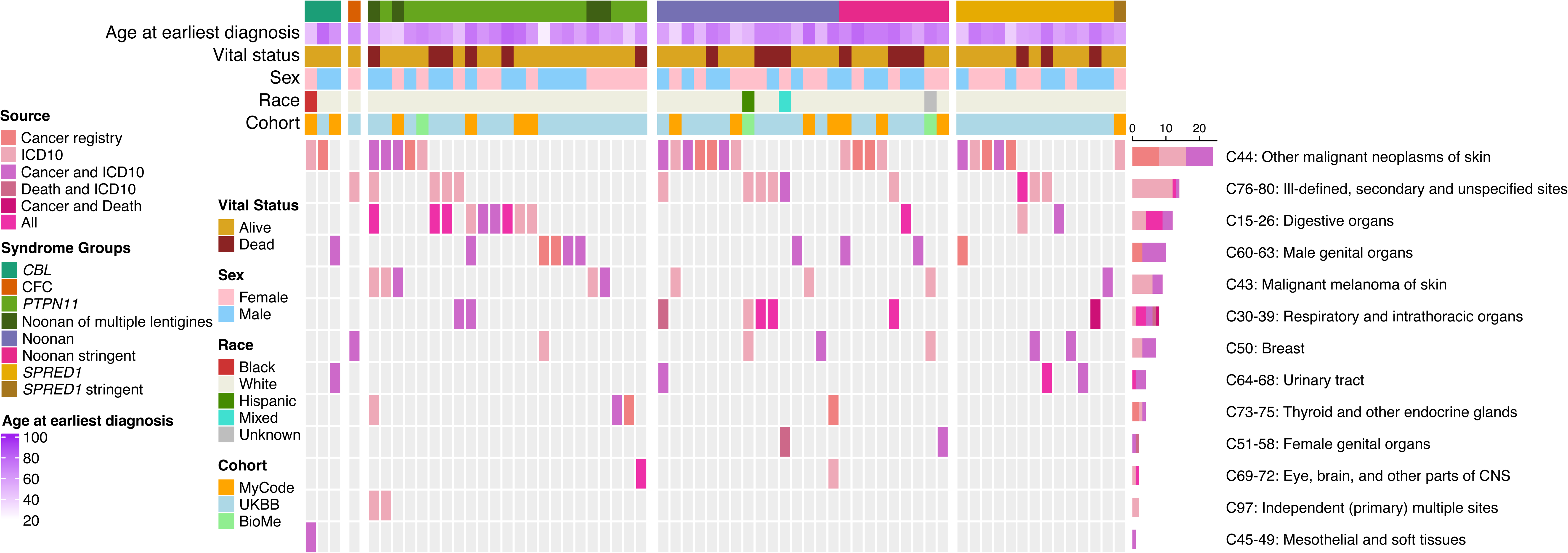
Oncoprint of cancer type in RASopathy heterozygotes in MyCode, UKBB and Bio*Me*.

### Cumulative cancer risk in RASopathy heterozygotes

Cumulative cancer risk was evaluated in RASopathy heterozygotes. This was only examined in NS and *SPRED1* heterozygotes in UKBB, since cancer cases reported in *CBL* and CFC heterozygotes were too few to provide meaningful analyses. Using Kaplan-Meier cumulative incidence, there was no difference in cumulative risk in NS heterozygotes in UKBB (Figure 3A**,B**), regardless of broad (Pcox=0.72) or stringent filtering criteria (Pcox= 0.85); similar findings were observed in MyCode (broad Pcox= 0.080; stringent Pcox= 0.069). (Figure 4 **A,B**) However, *SPRED1* heterozygotes showed significant differences (Pcox= 9.5×10^-4^) (Figure 3C), with cancer cumulative incidence increasing shortly before age 60. However, these finding could not be replicated in MyCode due to the lower number of *SPRED1-*heterozygotes.

**Figure 3.**
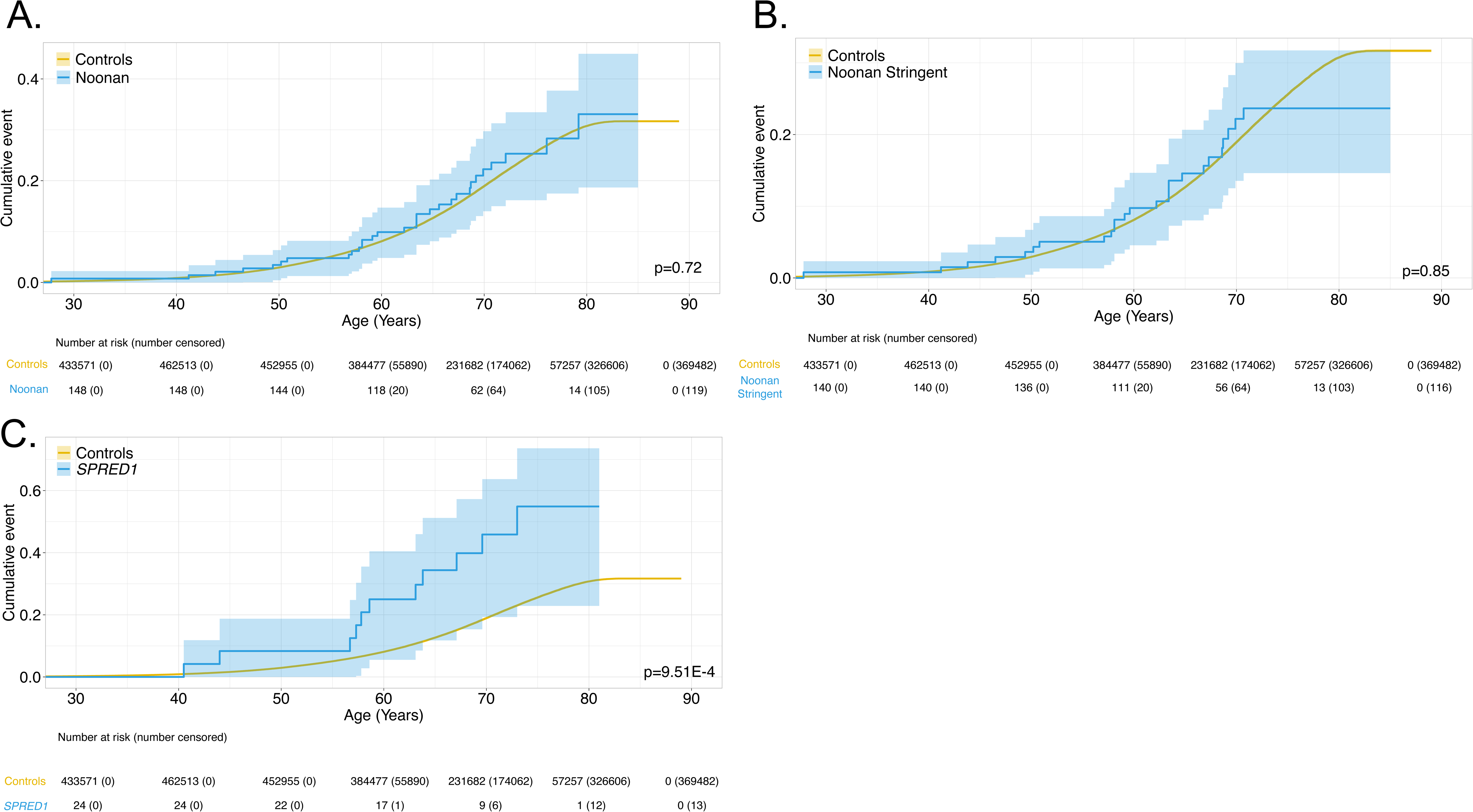
Time-dependent penetrance in Noonan-heterozygotes for cancer (**panel A**) and with stringent filtering (**panel B**) and in *SPRED1*-heterozygotes (**panel C**) in UKBB.

**Figure 4.**
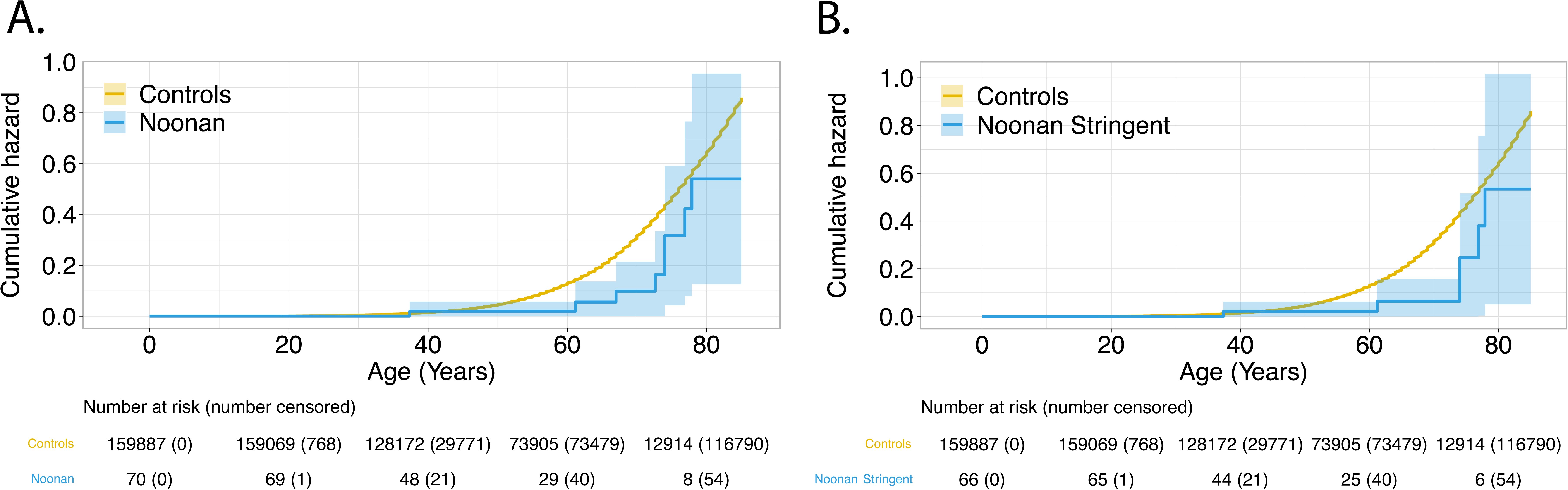
Time-dependent penetrance in Noonan-heterozygotes for cancer (**panel A**) and with stringent filtering (**panel B**) in MyCode.

### Overall survival in RASopathy heterozygotes

We next investigated if RASopathy heterozygotes were associated with increased mortality. There were < 5 deaths each in CFC-, *CBL*- and *SPRED1*-heterozygotes but 21 (14%) and 11 (15%) deaths in NS-heterozygotes in UKBB and MyCode, respectively. In MyCode, there was no significant difference in survival in NS-heterozygotes compared to controls (broad Pcox= 0.43, stringent Pcox= 0.63) (Figure 5A**,B**). In UKBB, NS-heterozygotes had less favorable survival compared to controls with a decrease in restricted mean age of about two years (Pcox= 0.0358; Figure 5C). However, when using stringent filtering criteria for NS-heterozygotes, no difference in survival was observed (Pcox= 0.127; Figure 5D). **Supplementary Figure 9** shows cause of death by ICD10 chapter grouping in UKBB; cancer was reported as the most common cause of mortality in RASopathy heterozygotes in UKBB, followed by cardiac and respiratory etiologies. However, when compared with controls, there were no significant differences in cancer and cardiac causes of death (p = 0.08 and 0.54, respectively), with a significant increase in a “respiratory system” cause of death (p = 0.02).

**Figure 5.**
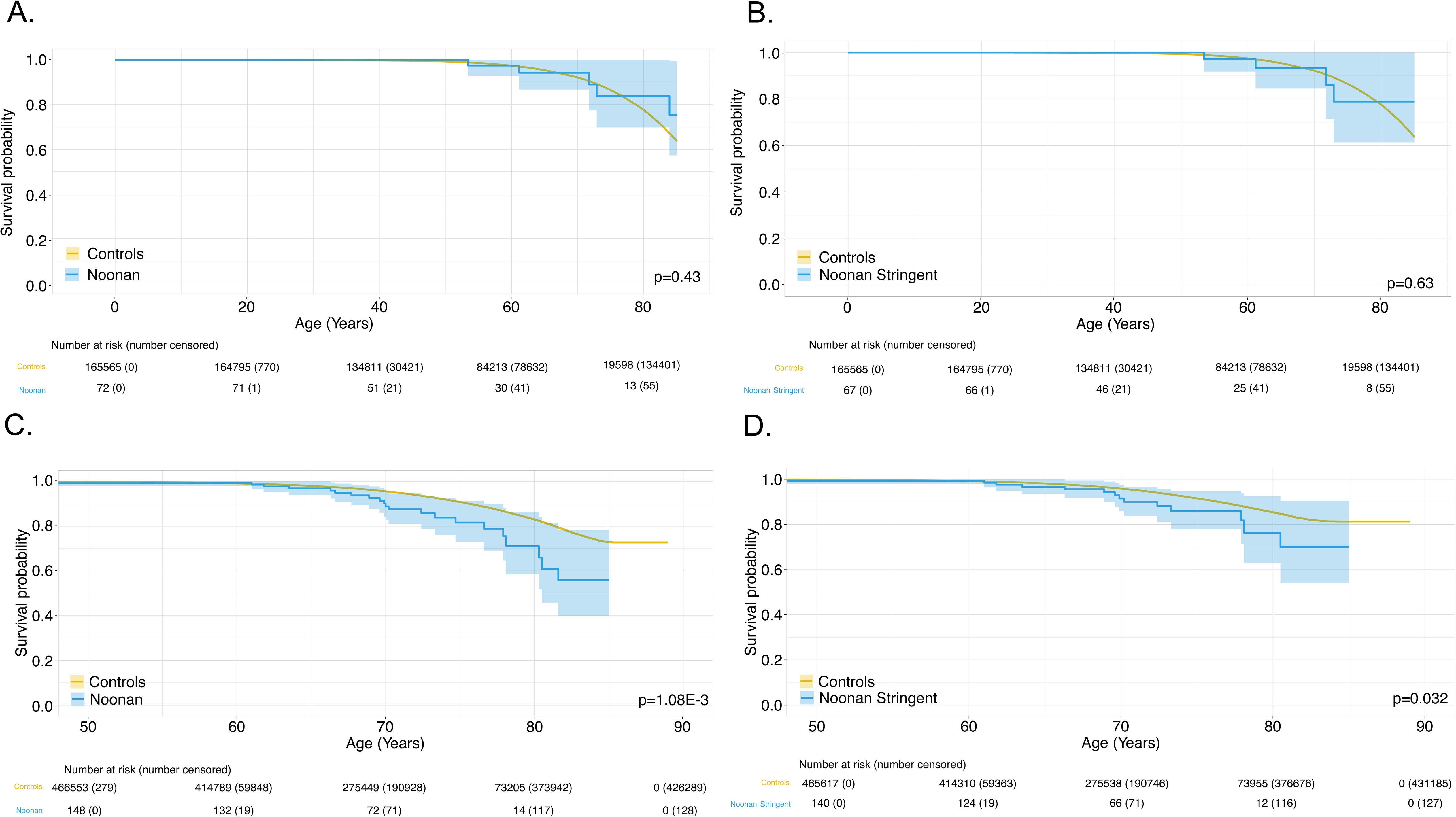
Time-dependent survival for Noonan-heterozygotes (**panel A**) and with stringent (**panel B**) filtering in MyCode; time-dependent survival for Noonan-heterozygotes (**panel C**) and with stringent (**panel D**) filtering in and UKBB.

## Discussion

In this investigation, genomic ascertainment of two population-based, exome-sequenced, EHR-linked cohorts were used to quantify Bonferroni-corrected risk of cancers arising from adults harboring P/LP germline variants in non-NF1 RASopathy genes. Notably, both cohorts had high power to detect elevated risk (OR>2) in NS heterozygotes for all but the rarest cancers. Genomic ascertainment quantifies risk based on genotype (not phenotype) and thus may reduce risk inflation arising from cancer ascertainment (case/family recruitment) by personal and/or family medical history. Notably, in contrast to numerous reports of children and adolescents with NS, where cancer rates are estimated at 4% by age 20, respectively^5,6^, there was no evidence of an increased cancer prevalence in adult NS heterozygotes in this analysis. Surprisingly, there was a significant excess of cancer risk in *SPRED1*-heterozygotes in UK Biobank but not MyCode. In both cohorts, CFC heterozygotes were more prevalent than previously reported; no *HRAS* P/LP variants were not detected in either cohort.

An increased risk of cancer was observed in *SPRED1*-heterozygotes in the UKBB with the elevated risk beginning in the 50s. The limited number (n=4) of *SPRED1*-heterozygotes in the MyCode cohort did not permit replication of this observation. While there have been some case reports about increased leukemia risk in Legius syndrome^25,26^, to our knowledge, this is the first comprehensive evaluation of cancer risk in this disorder. We acknowledge that the modest number of *SPRED1*-heterozygotes in both cohorts resulted in underpowered analysis and, thus, these findings merit replication in additional, diverse cohorts. For now, given that the excess cancer was driven by skin malignancies, it may be prudent to evaluate concerning skin findings in *SPRED1*-heterozygotes. Although there have been reports of increased cancer risk in *CBL* and CFC syndromes^27^, no significant excess cancer risk was observed in adult *CBL*- or CFC- heterozygotes. However, we acknowledge that the number of observations was small and that analyses of the cohorts were underpowered.

Notably, our study was well-powered to observe excess risk from non-rare cancers in NS heterozygotes. Although no excess cancer risk in adults or significantly different time-dependent penetrance of cancer was observed in NS-heterozygotes, one XY individual who was heterozygous for a P/LP variant in *PTPN11* had a history of a malignant neoplasm of the testis (at age 27.8) and breast cancer (at age 62) in the UKBB merits comment. This individual had no documented family history of breast cancer, history of hormone treatments, *BRCA1*, *BRCA2, PALB2* or *CHEK2* P/LP variants or other explanation on chart review. Despite the lack of evidence of a significant increase in cancer prevalence in NS-heterozygotes, the mean restricted age (a proxy for life expectancy) was reduced by about two years compared with controls in the UKBB, the only cohort in which this analysis was possible. However, when using “stringent” filtering, this difference diminished and was not statistically significant, and the two most common causes of death in NS heterozygotes (neoplasm, circulatory) were not significantly different from those of UKBB controls. Furthermore, while cancer risk in NSML is largely unknown, the four NSML heterozygotes with cancer all had been diagnosed with melanoma, suggesting a possible link between the increased skin findings in NSML and skin cancer risk. This will need to be carefully examined in additional cohorts to provide accurate recommendations for surveillance and management in NSML.

Clonal hematopoiesis (CH) is an age-associated process that arises from somatic mutations and clonal expansion in a hematopoietic precursor cell^28–30^. In addition to health consequences^30–35^, CH can confound identification of true germline variation and prevalence estimates^28,36–38^. Since many RASopathy genes are susceptible to CH^20^, we sequenced putative germline variants in archived, non-bloodline clinical tissue samples from the MyCode cohort. From these findings, we developed a conservative filtering strategy to reduce the likelihood of false-positive germline variation from CH. From this, we calculated the genomically ascertained prevalence of RASopathy heterozygotes (**Table 1**). Comparing these prevalences with published reports of phenotypically ascertained prevalence of RASopathy syndromes is tricky given the substantially different approaches to develop these estimates. In general, our estimates of the population prevalence of NS heterozygotes (those who may or may not have a phenotype) are less common that the with previous published reports of 1/1,000 – 1/2,500 for NS diagnoses^39^. There are likely many reasons for this, including a lack of formal clinical diagnostic criteria for NS, the “lumping” of other diagnoses (e.g., CFC) with NS before the widespread availability of genetic testing and ascertainment/survival biases in our cohorts. In contrast, our estimates of the prevalence of CFC heterozygotes are remarkably congruent and are much more common than published estimates of CFC syndrome (1/810,000)^40^. If replicated, CFC heterozygotes may be more frequent than currently estimated and that the clinical variability in CFC is much broader than previously appreciated. Prevalence from phenotypic ascertainment of Legius syndrome is reported to be 1/46,000–1/75,000^26,41–43^, which overlaps with our genomically ascertained estimate in MyCode. *SPRED1*-heterozygotes are more common in UKBB for unclear reasons. Given the expected rarity and severity of Costello syndrome, it was not surprising *HRAS*-heterozygotes were not observed in either cohort. To our knowledge, our estimates of *CBL*-heterozygotes prevalence are the first to be reported.

In other monogenic disorders, genomic ascertainment has shown greater prevalence of germline variation and reduced penetrance of phenotype than expected^44^. A previous investigation from our group using genomic ascertainment in the Bio*Me* and UKBB cohorts^4^ showed that NS- heterozygotes had significantly lower height z-scores compared to controls, a finding that was replicated in UKBB and MyCode in this study. Remarkably, no RASopathy-heterozygote in MyCode and only six in UKBB had the Q87 ICD-10 code in their EHR. This is evidence that the vast majority of NS-heterozygotes had not been previously diagnosed with a RASopathy, although they may have had mild relevant clinical features as suggested by their lower height z-scores. It is not clear if NS heterozygotes detected via genomic ascertainment represent incomplete penetrance of the syndrome or if these individuals harbor variants associated with more subtle clinical features and are therefore more likely to go undiagnosed. Nonetheless, proper detection and diagnosis may inform the medical management of these individuals.

Limitations of this investigation include the limited ancestral diversity of UKBB and MyCode. The UKBB dataset has been found to be have a “healthier volunteer” effect^45^, and the RASopathies are known to be associated with severe, early manifestations, including cardiac disease which can result in early death, and these individuals would therefore not be captured in older cohorts. Consequently, the penetrance and prevalence may not be fully representative of the broader general population. Furthermore, caution should still be used when interpreting prevalence, as common germline filtering criteria can still be contaminated by somatic variants, particularly in cases of CH. Ultimately, some cases will be excluded from this analysis (such as those resulting in early lethality) while contamination also occurs (due to CH, for example) and may help explain why our estimates are similar to reported prevalences which have been based on phenotypic ascertainment.

In summary, genomic ascertainment from two large biobanks did not show evidence of elevated cancer risk in adult NS-heterozygotes. However, there may be an increased cancer risk for adult NSML- and *SPRED1*-heterozygotes. RASopathies are notably under-diagnosed. CFC and *SPRED1* heterozygotes may be more common than previously appreciated. More research in broad and diverse cohorts is needed to replicate these findings that may inform future cancer screening guidelines for adults with RASopathies. While individuals ascertained through a genotype-first approach appear to have a milder phenotype, proper diagnosis may nevertheless inform optimal medical management and improve health in these individuals.

## Data Availability

All data produced are available online at https://www.ukbiobank.ac.uk/

## Acknowledgments

This work was supported by the Intramural Research program of the Division of Cancer Epidemiology and Genetics, National Cancer Institute, Rockville, MD. This research has been conducted using the UK Biobank Resource under Application Number 54389. This work used the computational resources of the NIH High Performance Computing Biowulf cluster. The authors are grateful to participants of the MyCode Community Initiative for use of their genomic clinical data, without whom parts of the study are not feasible. We thank the Geisinger-Regeneron Genetics DiscovEHR Collaboration for access to the genotype and phenotype data.

The content of this publication does not necessarily reflect the views or policies of the Department of Health and Human Services, nor does mention of trade names, commercial products or organizations imply endorsement by the U.S. Government.

## Notes

### Competing Interest Statement

The authors have declared no competing interest.

### Author Declarations

This study was approved by the Geisinger Institutional Review Board.

## References

1. Zenker M: Clinical overview on RASopathies. Am J Med Genet C Semin Med Genet 190:414–424, 2022

2. Rauen KA: Cardiofaciocutaneous Syndrome, in Adam MP, Mirzaa GM, Pagon RA, et al (eds): GeneReviews((R)). Seattle (WA), 1993

3. Tamburrino F, Scarano E, Schiavariello C, et al: Endocrinological manifestations in RASopathies. Am J Med Genet C Semin Med Genet 190:471–477, 2022

4. Wenger BM, Patel N, Lui M, et al: A genotype-first approach to exploring Mendelian cardiovascular traits with clear external manifestations. Genet Med 23:94–102, 2021

5. Kratz CP, Franke L, Peters H, et al: Cancer spectrum and frequency among children with Noonan, Costello, and cardio-facio-cutaneous syndromes. Br J Cancer 112:1392–7, 2015

6. Kratz CP, Rapisuwon S, Reed H, et al: Cancer in Noonan, Costello, cardiofaciocutaneous and LEOPARD syndromes. Am J Med Genet C Semin Med Genet 157C:83–9, 2011

7. Gripp KW, Morse LA, Axelrad M, et al: Costello syndrome: Clinical phenotype, genotype, and management guidelines. Am J Med Genet A 179:1725–1744, 2019

8. Astiazaran-Symonds E, Ney GM, Higgs C, et al: Cancer in Costello syndrome: a systematic review and meta-analysis. Br J Cancer 128:2089–2096, 2023

9. Ney G, Gross A, Livinski A, et al: Cancer incidence and surveillance strategies in individuals with RASopathies. Am J Med Genet C Semin Med Genet 190:530–540, 2022

10. Yang F, Long N, Anekpuritanang T, et al: Identification and prioritization of myeloid malignancy germline variants in a large cohort of adult patients with AML. Blood 139:1208–1221, 2022

11. Bycroft C, Freeman C, Petkova D, et al: The UK Biobank resource with deep phenotyping and genomic data. Nature 562:203–209, 2018

12. Dewey FE, Murray MF, Overton JD, et al: Distribution and clinical impact of functional variants in 50,726 whole-exome sequences from the DiscovEHR study. Science 354, 2016

13. Tayo BO, Teil M, Tong L, et al: Genetic background of patients from a university medical center in Manhattan: implications for personalized medicine. PLoS One 6:e19166, 2011

14. Grant AR, Cushman BJ, Cave H, et al: Assessing the gene-disease association of 19 genes with the RASopathies using the ClinGen gene curation framework. Hum Mutat 39:1485–1493, 2018

15. Cingolani P, Platts A, Wang le L, et al: A program for annotating and predicting the effects of single nucleotide polymorphisms, SnpEff: SNPs in the genome of Drosophila melanogaster strain w1118; iso-2; iso-3. Fly (Austin) 6:80–92, 2012

16. Li Q, Wang K: InterVar: Clinical Interpretation of Genetic Variants by the 2015 ACMG-AMP Guidelines. Am J Hum Genet 100:267–280, 2017

17. Kim J, Gianferante M, Karyadi DM, et al: Frequency of Pathogenic Germline Variants in Cancer-Susceptibility Genes in the Childhood Cancer Survivor Study. JNCI Cancer Spectr 5, 2021

18. Kim J, Vaksman Z, Egolf LE, et al: Germline pathogenic variants in neuroblastoma patients are enriched in BARD1 and predict worse survival. J Natl Cancer Inst 116:149–159, 2024

19. Martincorena I: Somatic mutation and clonal expansions in human tissues. Genome Med 11:35, 2019

20. Sperling AS, Gibson CJ, Ebert BL: The genetics of myelodysplastic syndrome: from clonal haematopoiesis to secondary leukaemia. Nat Rev Cancer 17:5–19, 2017

21. Chow S-C, Shao J, Wang H, et al: Sample size calculations in clinical research (ed Third edition.). Boca Raton, Taylor & Francis, 2017

22. Mendez HM, Opitz JM: Noonan syndrome: a review. Am J Med Genet 21:493–506, 1985

23. Roberts AE, Adam MP, Mirzaa GM, et al: Noonan Syndrome. GeneReviews(®), 1993

24. Legius E, Stevenson D, Adam MP, et al: Legius Syndrome. GeneReviews(®), 1993

25. Pasmant E, Sabbagh A, Hanna N, et al: SPRED1 germline mutations caused a neurofibromatosis type 1 overlapping phenotype. J Med Genet 46:425–30, 2009

26. Pasmant E, Gilbert-Dussardier B, Petit A, et al: SPRED1, a RAS MAPK pathway inhibitor that causes Legius syndrome, is a tumour suppressor downregulated in paediatric acute myeloblastic leukaemia. Oncogene 34:631–8, 2015

27. Gross AM, Frone M, Gripp KW, et al: Advancing RAS/RASopathy therapies: An NCI-sponsored intramural and extramural collaboration for the study of RASopathies. Am J Med Genet A 182:866–876, 2020

28. Kar SP, Quiros PM, Gu M, et al: Genome-wide analyses of 200,453 individuals yield new insights into the causes and consequences of clonal hematopoiesis. Nat Genet 54:1155–1166, 2022

29. Jaiswal S, Ebert BL: Clonal hematopoiesis in human aging and disease. Science 366, 2019

30. Xie M, Lu C, Wang J, et al: Age-related mutations associated with clonal hematopoietic expansion and malignancies. Nat Med 20:1472–8, 2014

31. Jaiswal S, Fontanillas P, Flannick J, et al: Age-related clonal hematopoiesis associated with adverse outcomes. N Engl J Med 371:2488–98, 2014

32. Jaiswal S, Natarajan P, Silver AJ, et al: Clonal Hematopoiesis and Risk of Atherosclerotic Cardiovascular Disease. N Engl J Med 377:111–121, 2017

33. Genovese G, Kahler AK, Handsaker RE, et al: Clonal hematopoiesis and blood-cancer risk inferred from blood DNA sequence. N Engl J Med 371:2477–87, 2014

34. Buttigieg MM, Rauh MJ: Clonal Hematopoiesis: Updates and Implications at the Solid Tumor-Immune Interface. JCO Precis Oncol 7:e2300132, 2023

35. Bolton KL, Gillis NK, Coombs CC, et al: Managing Clonal Hematopoiesis in Patients With Solid Tumors. J Clin Oncol 37:7–11, 2019

36. Brunet T, Berutti R, Dill V, et al: Clonal hematopoiesis as a pitfall in germline variant interpretation in the context of Mendelian disorders. Hum Mol Genet 31:2386–2395, 2022

37. Mester JL, Jackson SA, Postula K, et al: Apparently Heterozygous TP53 Pathogenic Variants May Be Blood Limited in Patients Undergoing Hereditary Cancer Panel Testing. J Mol Diagn 22:396–404, 2020

38. Batalini F, Peacock EG, Stobie L, et al: Li-Fraumeni syndrome: not a straightforward diagnosis anymore-the interpretation of pathogenic variants of low allele frequency and the differences between germline PVs, mosaicism, and clonal hematopoiesis. Breast Cancer Res 21:107, 2019

39. Roberts AE: Noonan Syndrome, in Adam MP, Feldman J, Mirzaa GM, et al (eds): GeneReviews((R)). Seattle (WA), 1993

40. Abe Y, Aoki Y, Kuriyama S, et al: Prevalence and clinical features of Costello syndrome and cardio-facio-cutaneous syndrome in Japan: findings from a nationwide epidemiological survey. Am J Med Genet A 158A:1083–94, 2012

41. Messiaen L, Yao S, Brems H, et al: Clinical and mutational spectrum of neurofibromatosis type 1-like syndrome. JAMA 302:2111–8, 2009

42. Evans DG, Bowers N, Burkitt-Wright E, et al: Comprehensive RNA Analysis of the NF1 Gene in Classically Affected NF1 Affected Individuals Meeting NIH Criteria has High Sensitivity and Mutation Negative Testing is Reassuring in Isolated Cases With Pigmentary Features Only. EBioMedicine 7:212–20, 2016

43. Giugliano T, Santoro C, Torella A, et al: Clinical and Genetic Findings in Children with Neurofibromatosis Type 1, Legius Syndrome, and Other Related Neurocutaneous Disorders. Genes (Basel) 10, 2019

44. Mirshahi UL, Colclough K, Wright CF, et al: Reduced penetrance of MODY-associated HNF1A/HNF4A variants but not GCK variants in clinically unselected cohorts. Am J Hum Genet 109:2018–2028, 2022

45. Fry A, Littlejohns TJ, Sudlow C, et al: Comparison of Sociodemographic and Health-Related Characteristics of UK Biobank Participants With Those of the General Population. Am J Epidemiol 186:1026–1034, 2017

