## Supplementary figures and images for "Genomic ascertainment to quantify prevalence and cancer risk in adults with pathogenic and likely pathogenic germline variants in RASopathy genes"

### Supplemental Figure 1

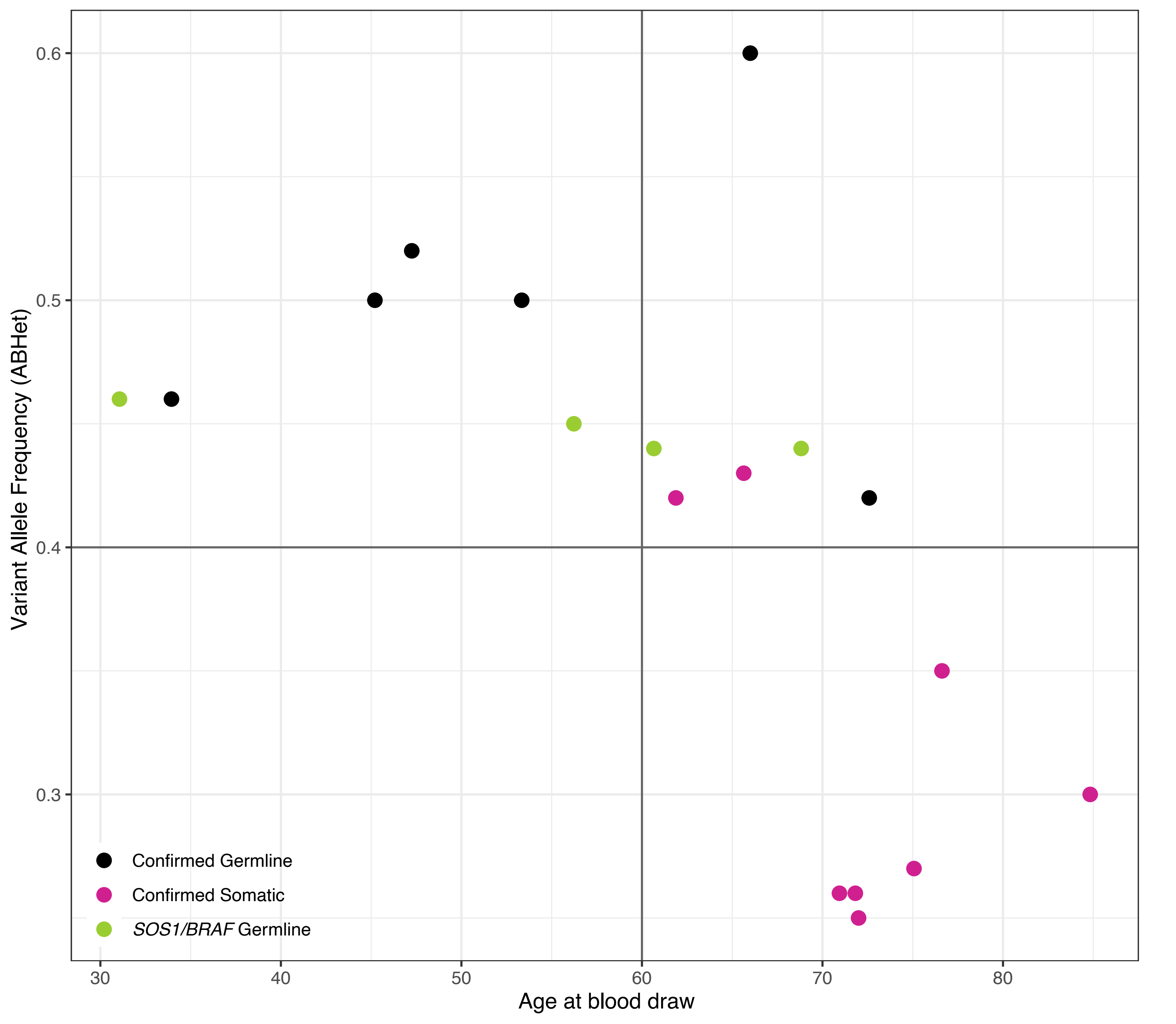

### Supplemental Figure 2

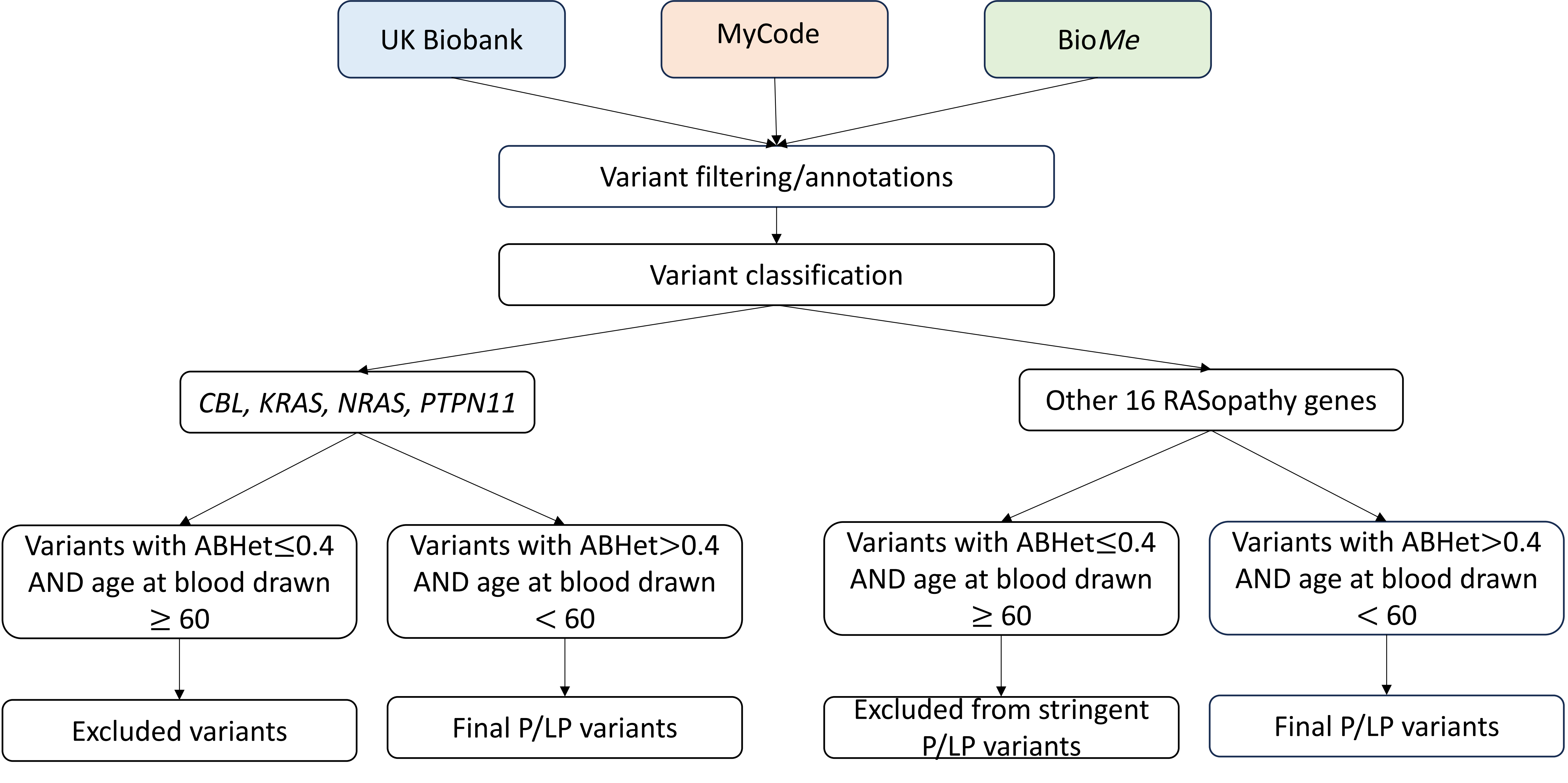

### Supplemental Figure 3

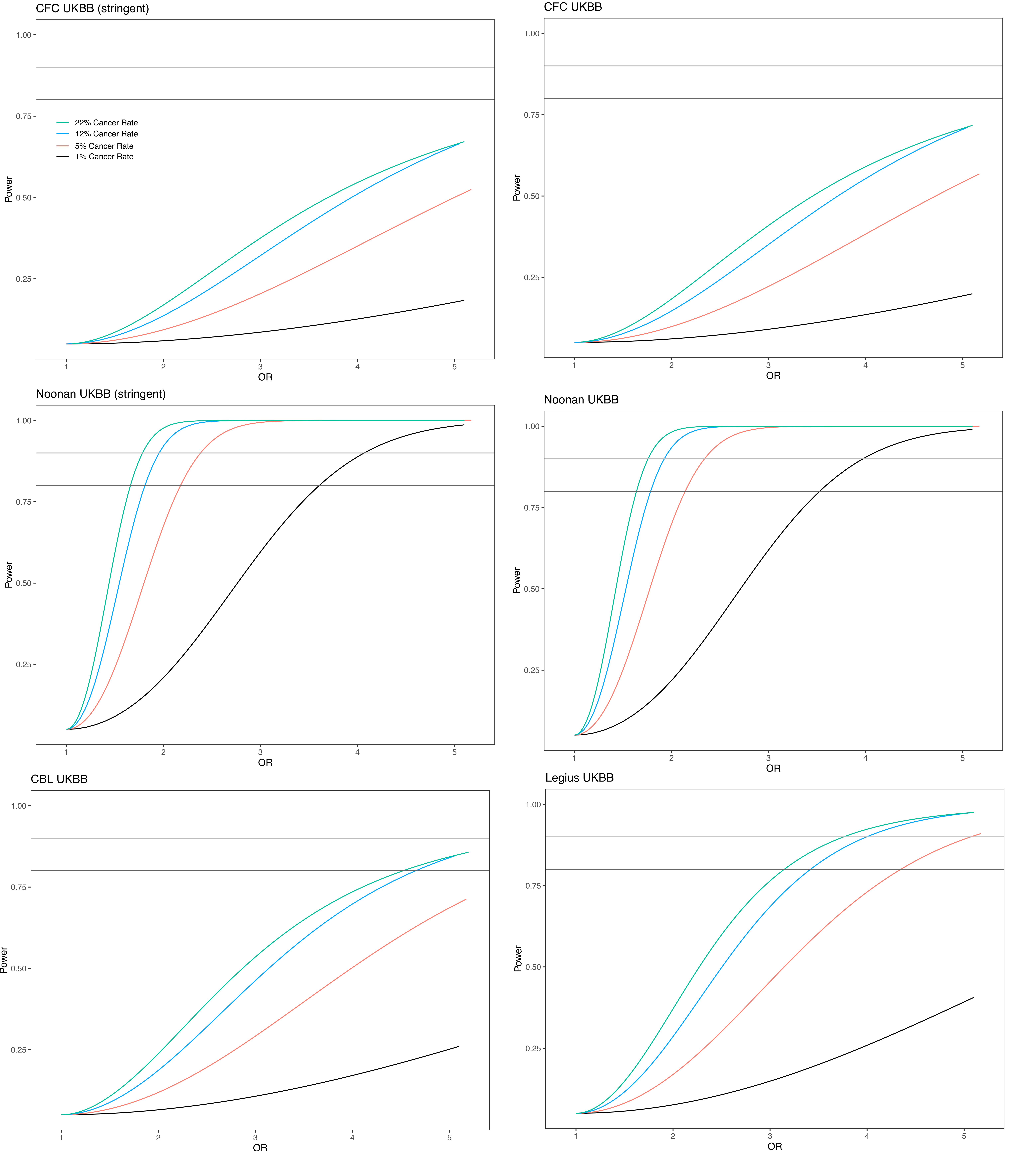

### Supplemental Figure 4

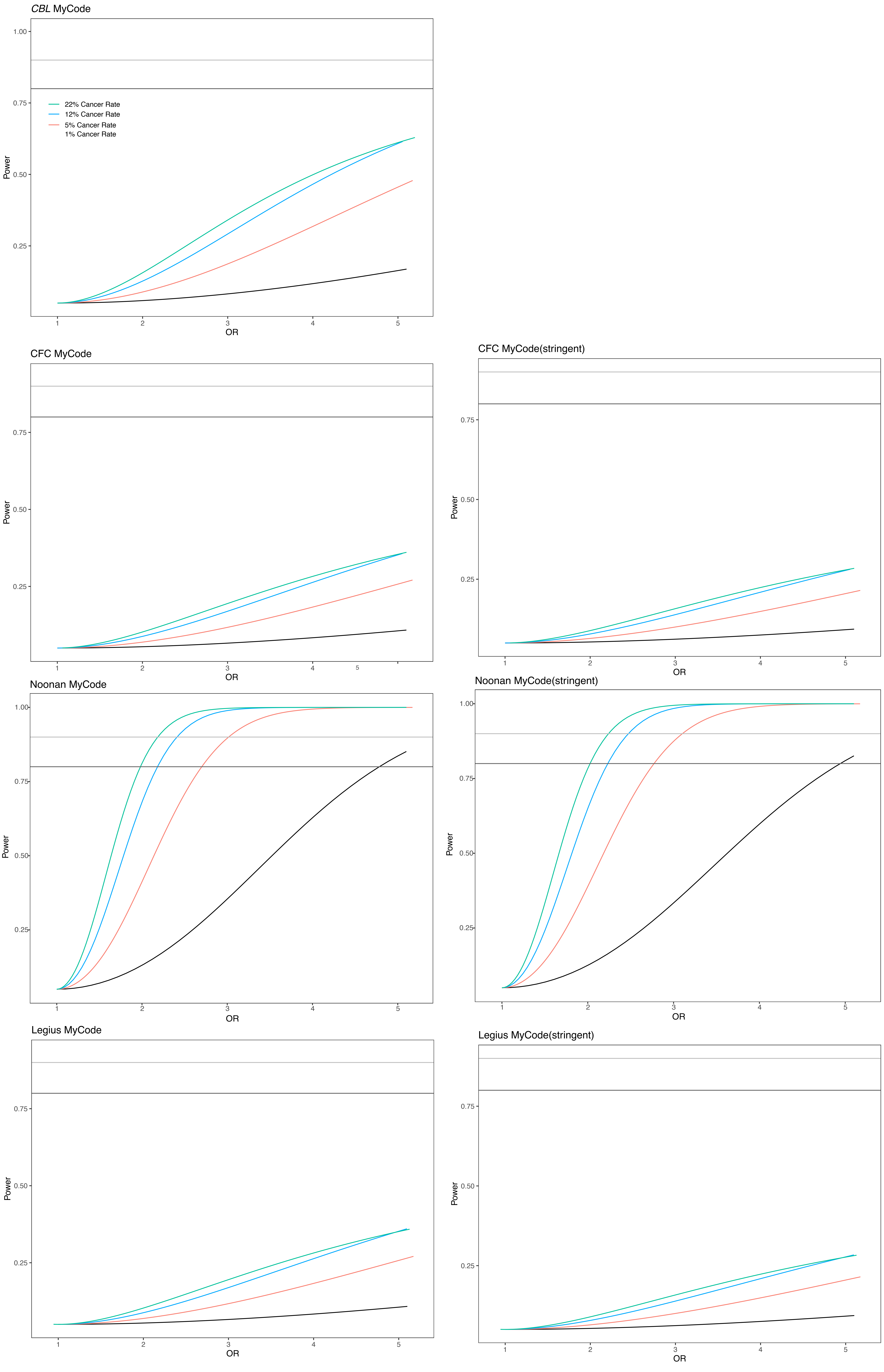

### Supplemental Figure 5

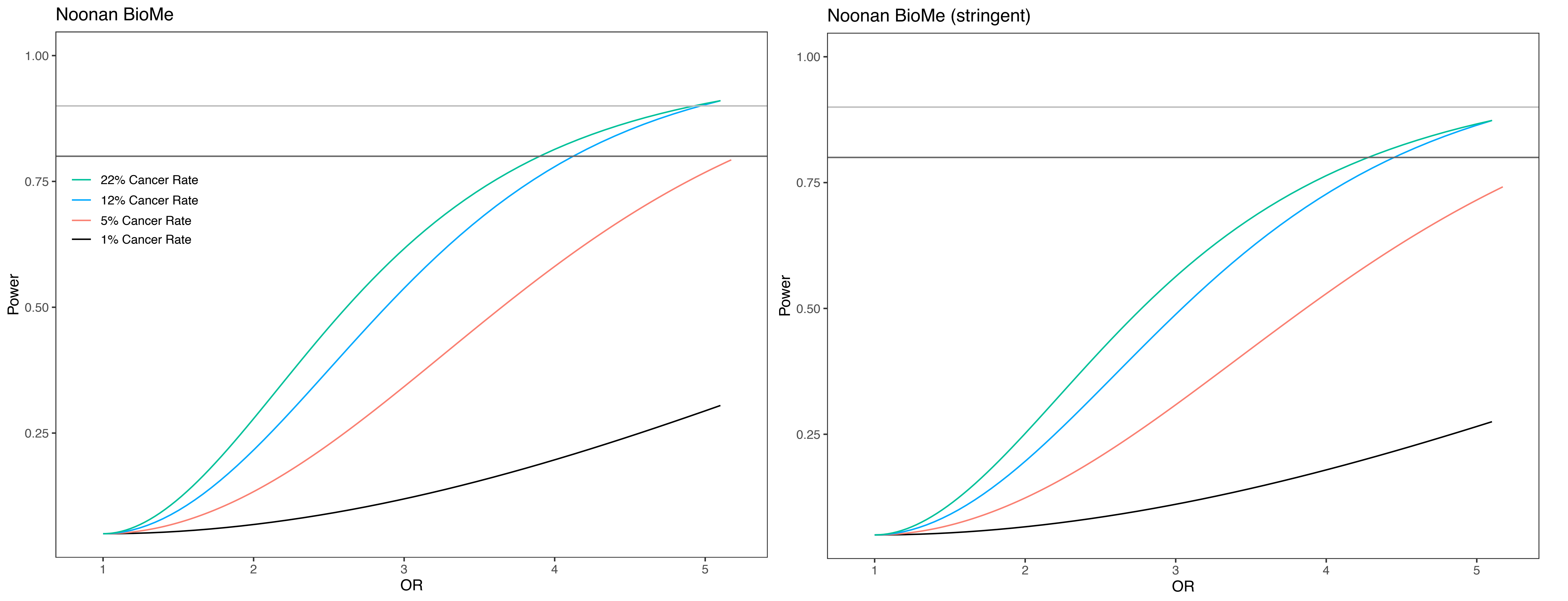

### Supplemental Figure 6

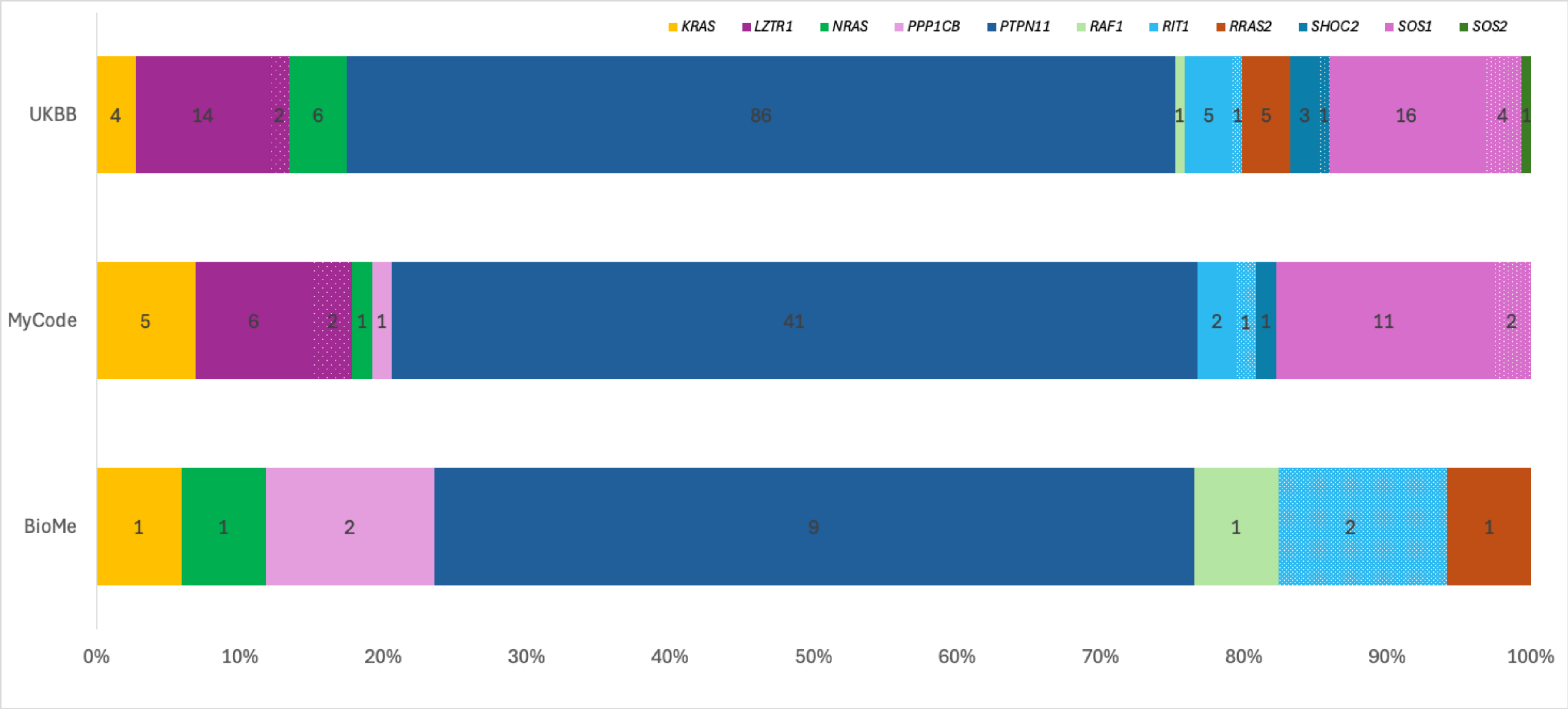

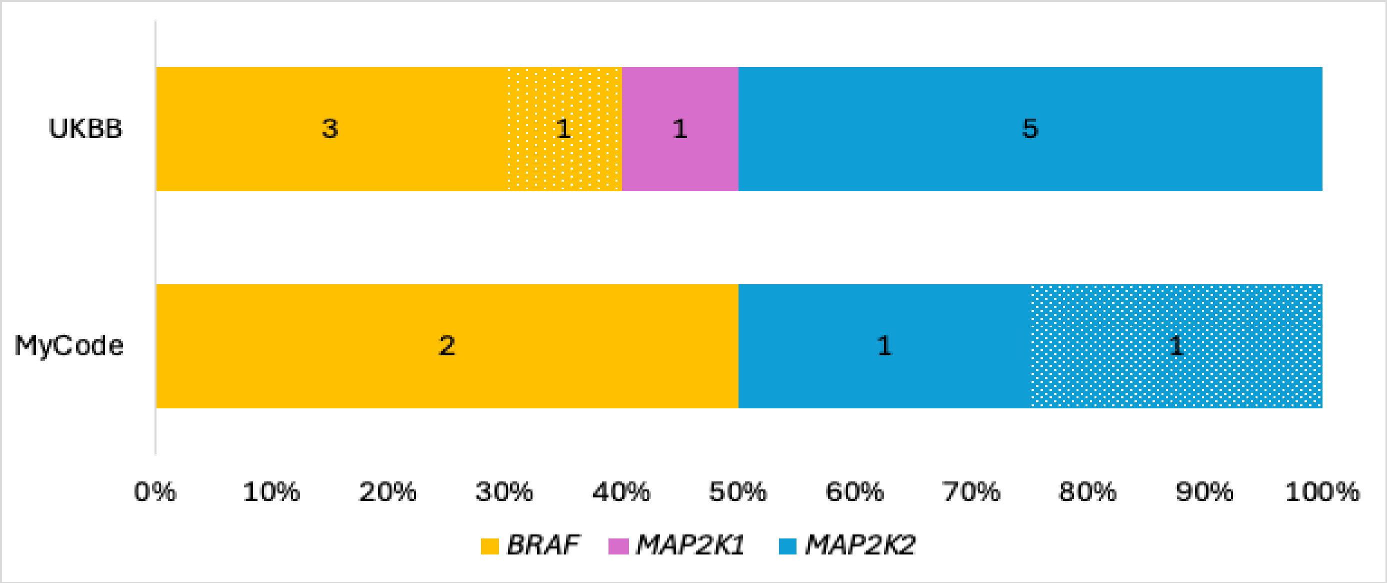

### Supplemental Figure 7

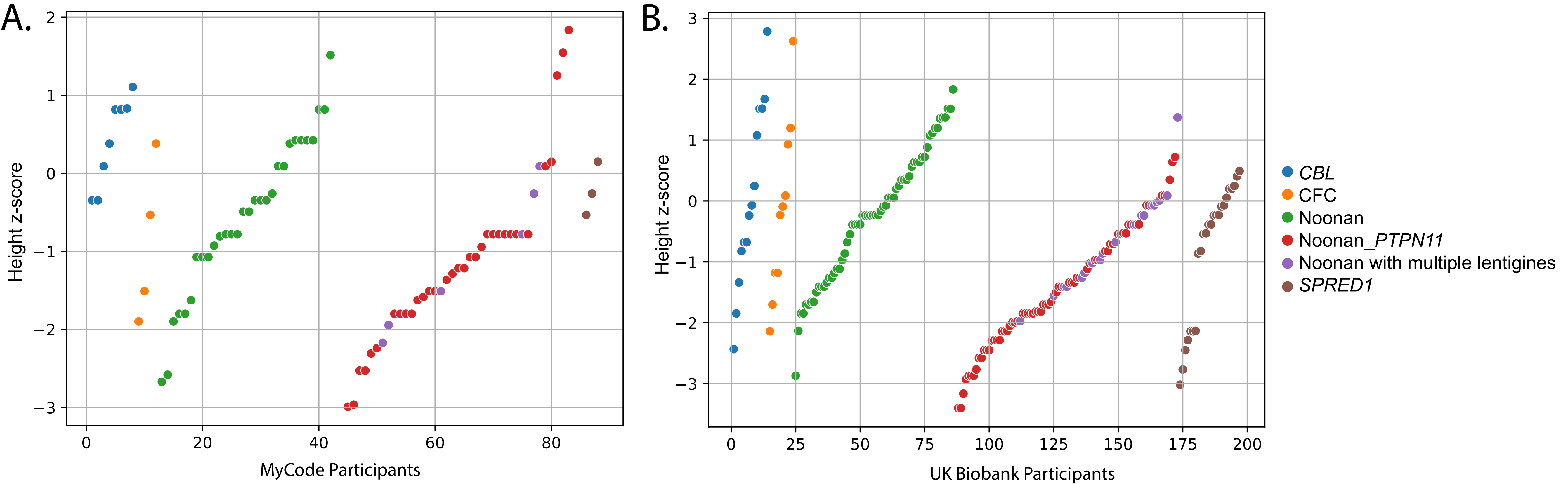

### Supplemental Figure 9

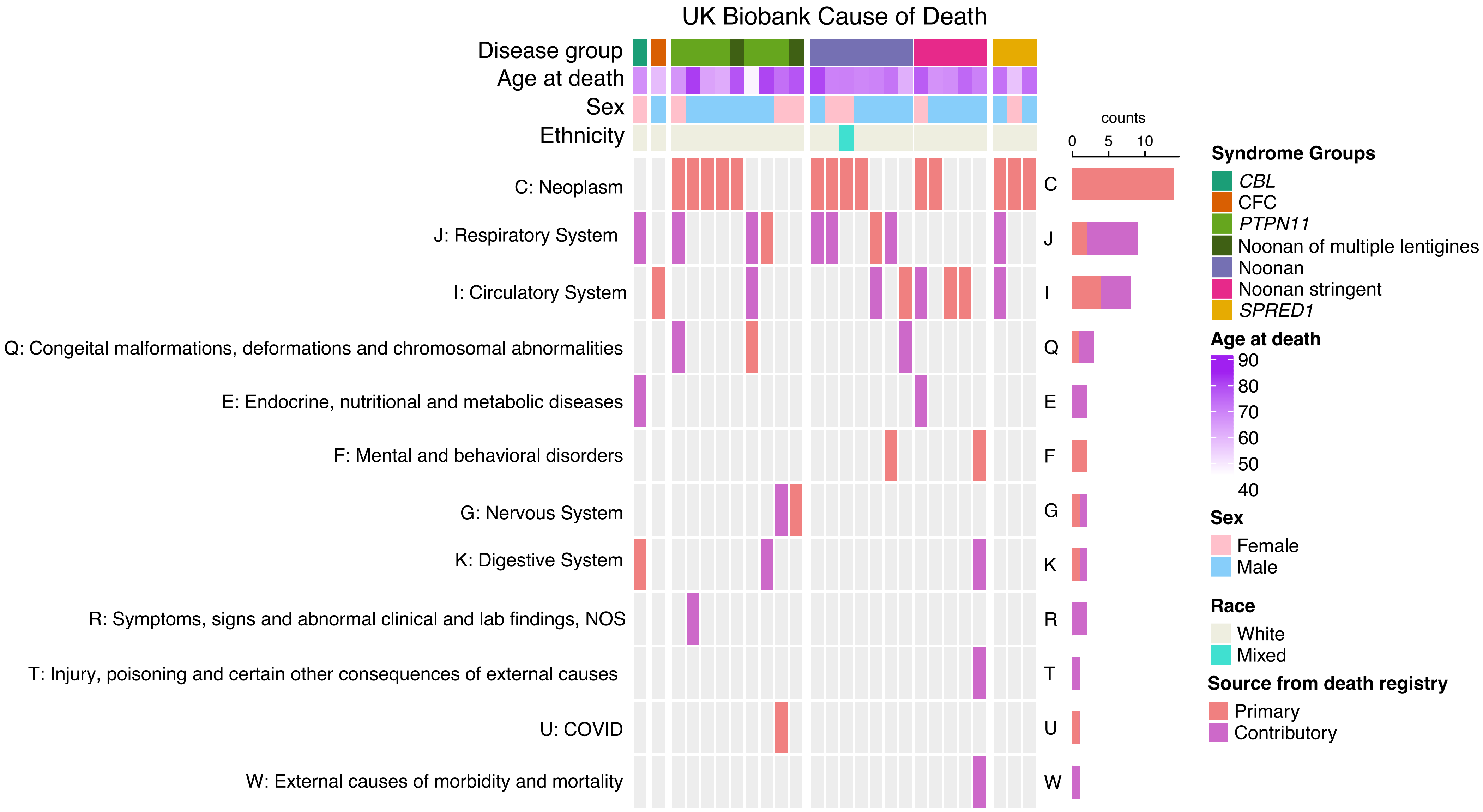
