## Supplemental Figure 8 for "Genomic ascertainment to quantify prevalence and cancer risk in adults with pathogenic and likely pathogenic germline variants in RASopathy genes"

Supplementary Figure 8: Lollipop plots of A)*BRAF* B)*CBL* C)*KRAS* D)*LZTR1* E)*MAP2K1* F)*MAP2K2* G)*NRAS* H)*PPP1CB* I)*PTPN11* J) *RAF1* K)*RIT1* L)*RRAS2* M)*SHOC2* N)*SOS1* O)*SOS2* P)*SPRED1*

**A.**

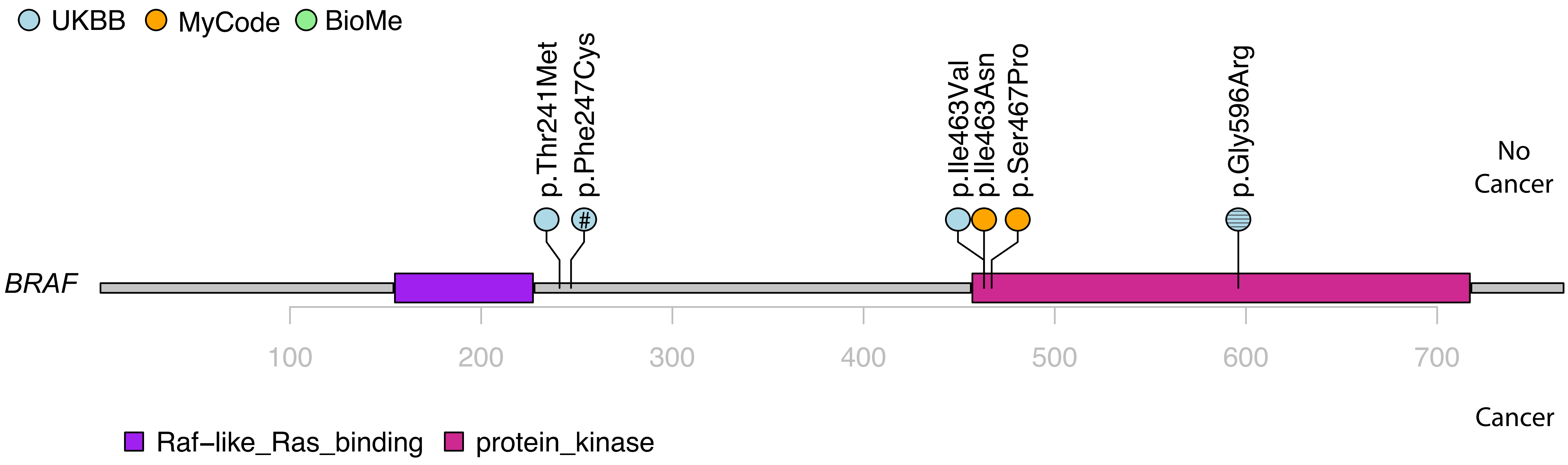


lines denotes variant that is dropped for stringent filter
### denotes death

**B.**

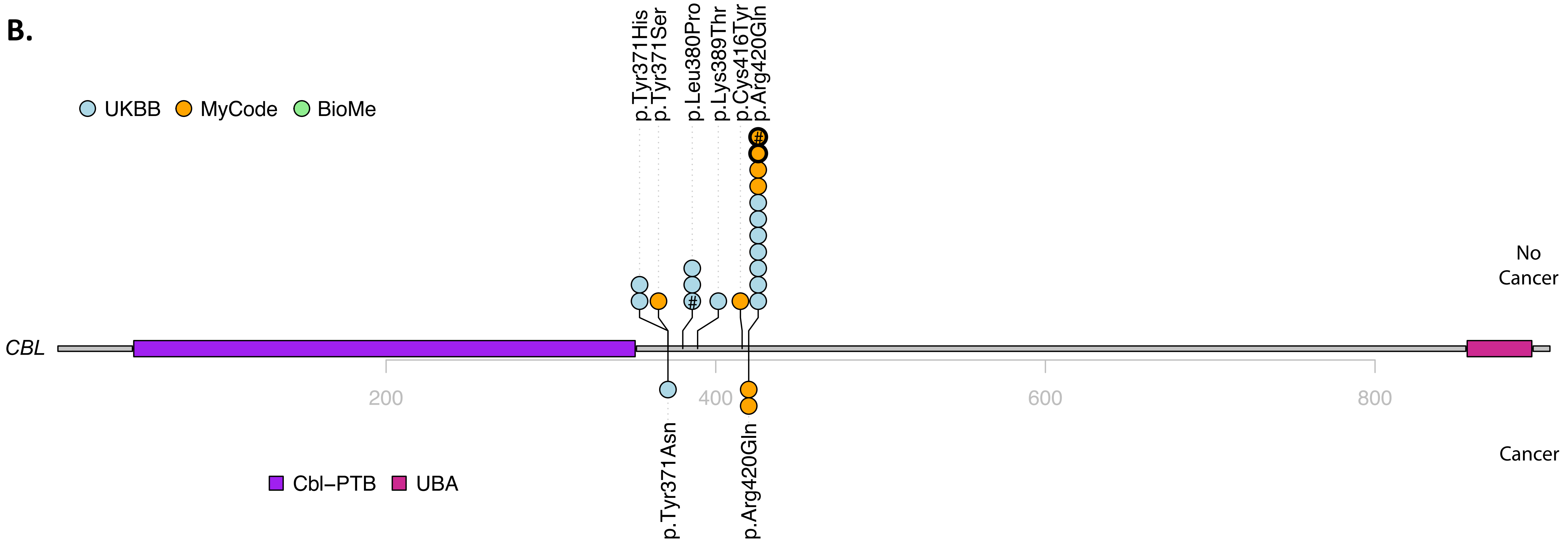


| | HGVS p. | Cancer | Histology | Age at diagnosis | Database |
|---|---|---|---|---|---|
| 1 | p.Tyr371Asn | C44.9 Malignant neoplasm of skin | Basal cell carcinoma, NOS | 78.4 | UKBB |
| 2 | p.Arg420Gln | C49.6 Malignant neoplasm of connective and soft tissue of head, face and neck | Dermatofibrosarcoma protuberans, fibrosarcomatous | 45 | MyCode |
| 3 | p.Arg420Gln | C61.9 Malignant neoplasm of prostate | Adenocarcinoma, NOS | 63 | MyCode |
| 3 | p.Arg420Gln | C67.9 Malignant neoplasm of trigone of bladder | Papillary transitional cell carcinoma, non-invasive | 77 | MyCode |

### denotes death
Bold outline represent related individuals

C.

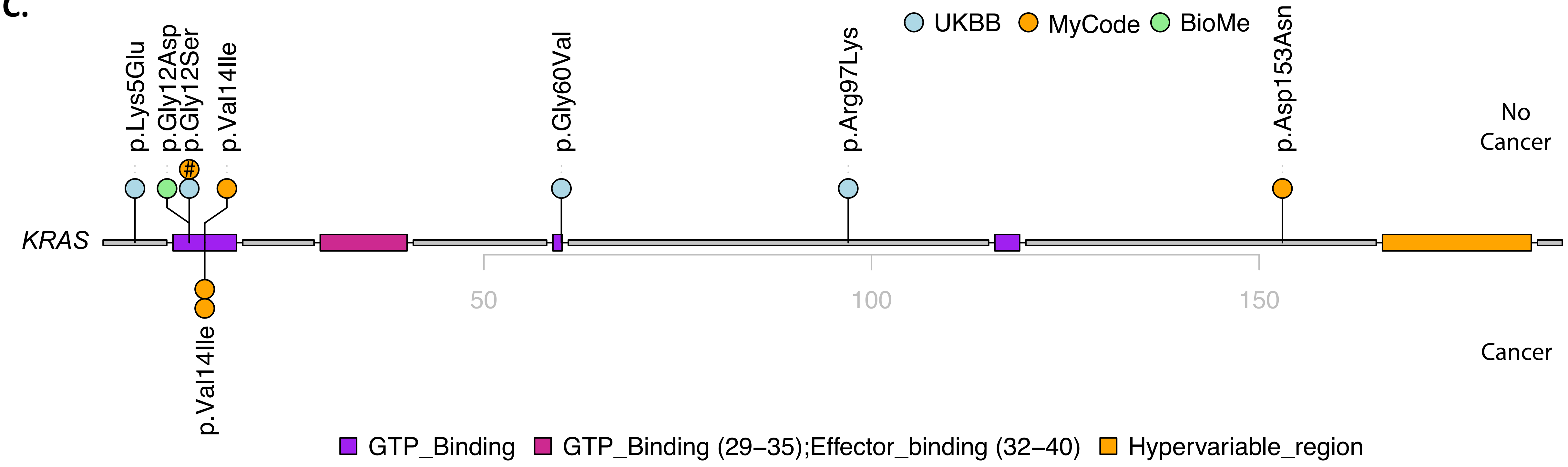


| | HGVS p. | Cancer | Histology | Age at diagnosis | Database |
|---|---|---|---|---|---|
| 1 | p.Val14Ile | C44.3 Other specified malignant neoplasm of skin of unspecified lower limb, including hip | . | 37.3 | MyCode |
| 1 | | C43.7 Malignant melanoma of unspecified lower limb, including hip | . | 46 | MyCode |
| 2 | p.Val14Ile | C44.3 Basal cell carcinoma of skin of nose | . | 77.9 | MyCode |

**D.**

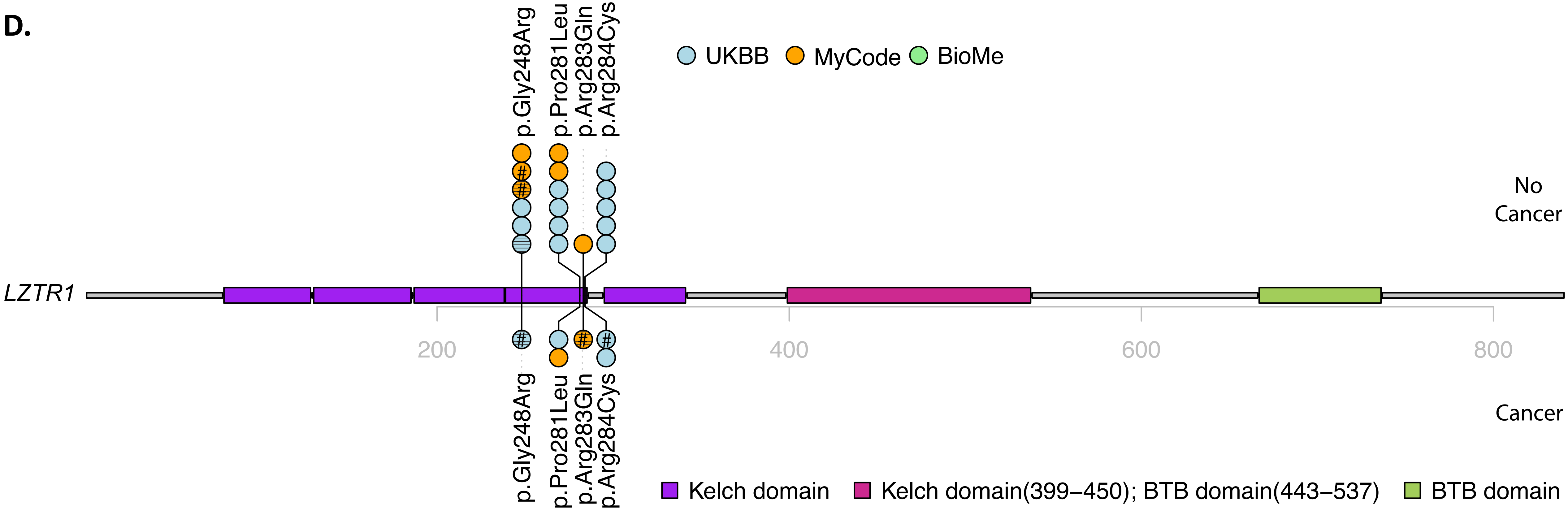


| | HGVS p. | Cancer | Histology | Age at diagnosis | Database |
|---|---|---|---|---|---|
| 1 | p.Gly248Arg | C61 Malignant neoplasm of prostate | Adenocarcinoma, NOS | 56.8 | UKBB |
| 2 | p.Pro281Leu | C62.9 Testis, unspecified | Seminoma, NOS | 50.2 | UKBB |
| 3 | p.Pro281Leu | C75.1 Malignant neoplasm of pineal gland | adenoma, NOS | 74 | MyCode |
| 3 | | C72.5Malignant neoplasm of unspecified cranial nerve | . | 76 | MyCode |
| 4 | p.Arg283Gln | C61.9 Malignant neoplasm of prostate | Adenocarcinoma, NOS | 67 | MyCode |
| 4 | | C44 Basal cell carcinoma | . | 85 | MyCode |
| 5 | p.Arg284Cys | C44.9 Malignant neoplasm of skin, unspecified; C44.2 Skin of ear and external auricular canal | Basal cell carcinoma, NOS | 70.7 | UKBB |
| 6 | p.Arg284Cys | C67.9 Bladder, unspecified | Transitional cell carcinoma, NOS | 58.1 | UKBB |
| 6 | | C44.4 Skin of scalp and neck | Basal cell carcinoma, NOS | 80.5 | UKBB |
| 6 | | C34.9 Bronchus or lung, unspecified; C34.1 Upper lobe, bronchus or lung | . | 81.6 | UKBB |
| 6 | | C79.9 Secondary malignant neoplasm, unspecified site | . | 81 | UKBB |

E.

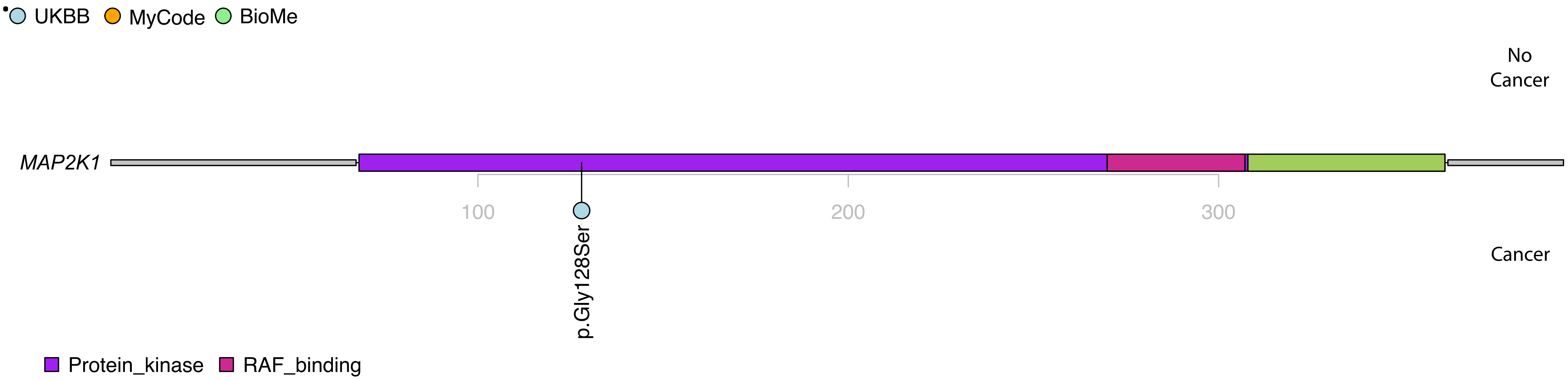


| | HGVS p. | Cancer | Histology | Age at diagnosis | Database |
|---|---|---|---|---|---|
| 1 | p.Gly128Ser | C50.9 Breast, unspecified; C50.3 Lower-inner quadrant of breast | Infiltrating duct carcinoma, NOS | 65.8 | UKBB |
| | | C77.3 Axillary and upper limb lymph nodes | . | 66 | UKBB |

F.

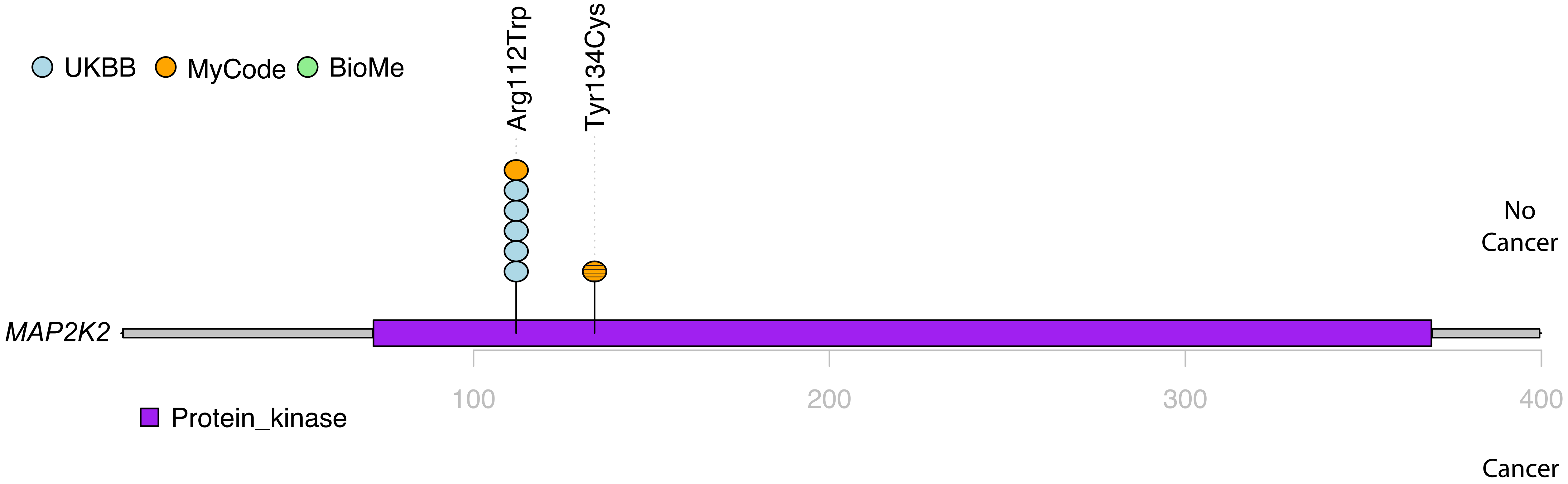

UKBB
MyCode
BioMe
Arg112Trp
Tyr134Cys
No
Cancer
MAP2K2
100
200
300
400
Protein_kinase
Cancer

**G.**

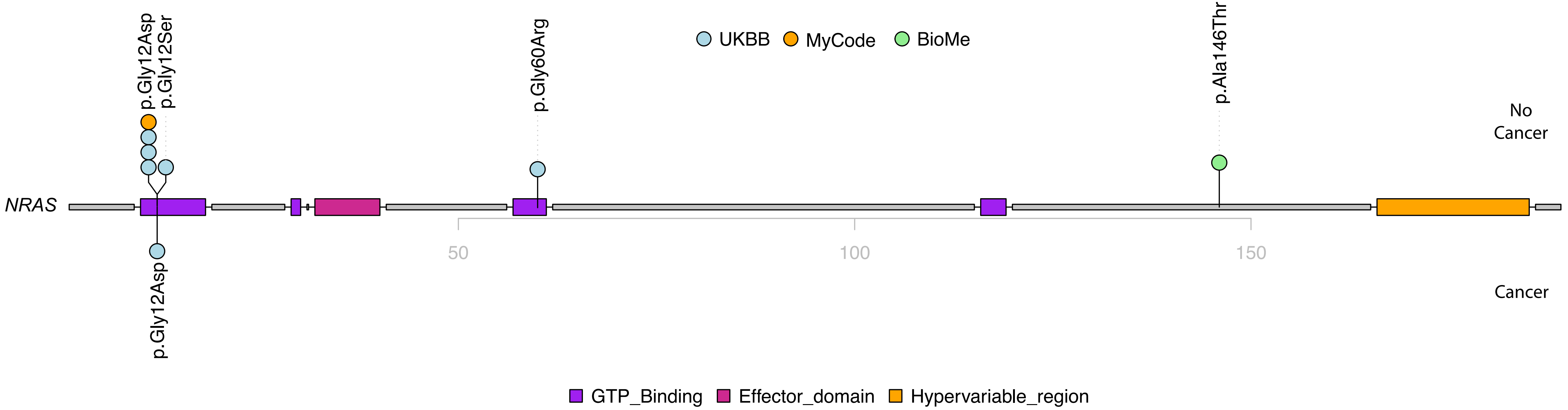


| | HGVS p. | Cancer | Histology | Age at diagnosis | Database |
|---|---|---|---|---|---|
| 1 | p.Gly12Asp | C44.3 Skin of other and unspecified parts of face | Basal cell carcinoma, NOS | 49.4 | UKBB |

**H.**

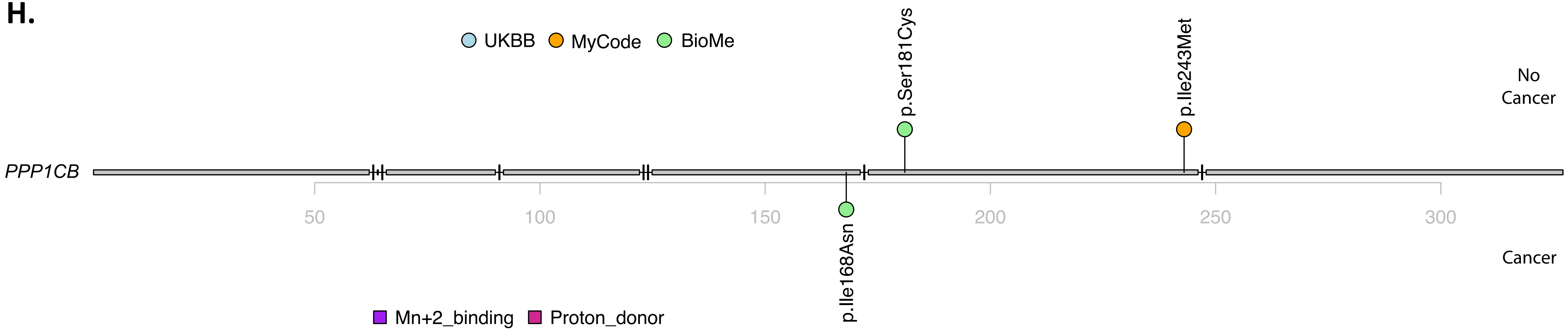


| | HGVS p. | Cancer | Histology | Age at diagnosis | Database |
|---|---|---|---|---|---|
| 1 | p.Ile168Asn | C50.9 Malignant neoplasm of unspecified site of unspecified female breast | . | 50 | BioMe |
| 1 | | C34.9 Malignant neoplasm of unspecified part of unspecified bronchus or lung | . | 55 | BioMe |

I.

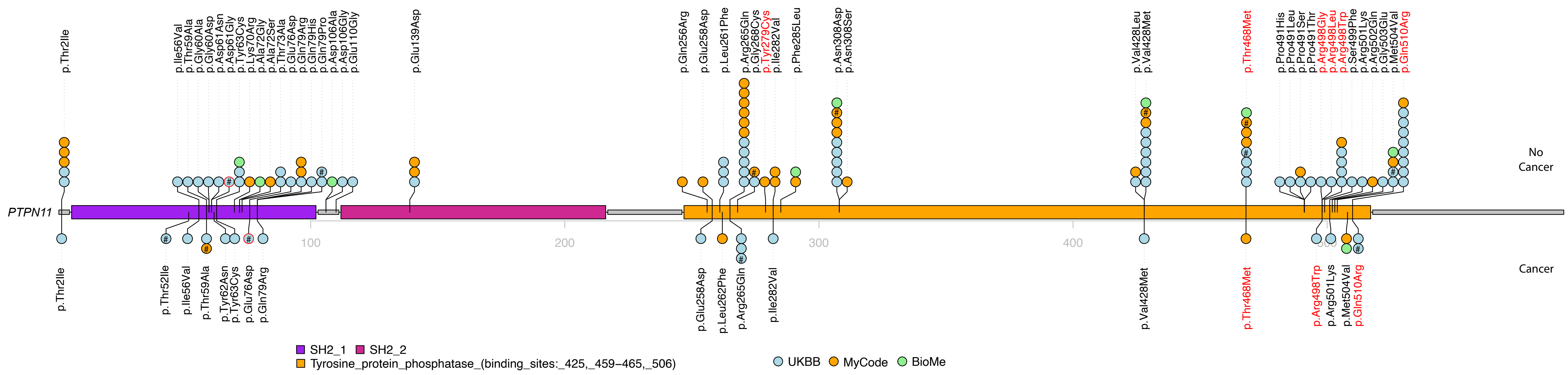
PTPN11
No Cancer
Cancer
100
200
300
400
SH2_1
SH2_2
Tyrosine_protein_phosphatase_(binding_sites:_425,_459–465,_506)
UKBB
MyCode
BioMe
p.Thr2Ile
p.Ile56Val
p.Thr59Ala
p.Gly60Ala
p.Gly60Asp
p.Asp61Asn
p.Asp61Gly
p.Tyr63Cys
p.Lys70Arg
p.Ala72Gly
p.Ala72Ser
p.Thr73Ala
p.Glu76Asp
p.Gln79Arg
p.Gln79His
p.Gln79Pro
p.Asp106Ala
p.Asp106Gly
p.Glu110Gly
p.Glu139Asp
p.Gln256Arg
p.Glu258Asp
p.Leu261Phe
p.Arg265Gln
p.Gly268Cys
p.Tyr279Cys
p.Ile282Val
p.Phe285Leu
p.Asn308Asp
p.Asn308Ser
p.Val428Leu
p.Val428Met
p.Thr468Met
p.Pro491His
p.Pro491Leu
p.Pro491Ser
p.Pro491Thr
p.Arg498Gly
p.Arg498Leu
p.Arg498Trp
p.Ser499Phe
p.Arg501Lys
p.Arg502Gln
p.Gly503Glu
p.Met504Val
p.Gln510Arg
p.Thr52Ile
p.Tyr62Asn
p.Leu262Phe

| | HGVS p. | Cancer | Histology | Age at diagnosis | Database |
|---|---|---|---|---|---|
| 1 | p.Thr2Ile | C73 Malignant neoplasm of thyroid gland | Papillary adenocarcinoma, NOS | 59.1 | UKBB |
| 2 | p.Thr52Ile | C18.0 Caecum | Neoplasm, malignant | 80.1 | UKBB |
| 3 | p.Ile56Val | C16.2 Body of stomach | . | 63.4 | UKBB |
| 3 | | C79.5 Secondry malignant neoplasm of bone and bone marrow | . | 64 | UKBB |
| 4 | p.Thr59Ala | C61 Malignant neoplasm of prostate | Adenocarcinoma, NOS | 63.4 | UKBB |
| 5 | p.Thr59Ala | C34.1 Malignant neoplasm of unspecified part of unspecified bronchus or lung | Squamous cell carcinoma, NOS | 83 | MyCode |
| 5 | | C61.9 Malignant neoplasm of prostate | Adenocarcinoma, NOS | 74 | MyCode |
| 6 | p.Tyr62Asn | C20 Malignant neoplasm of rectum | Adenocarcinoma, NOS | 62.2 | UKBB |
| 7 | p.Tyr63Cys | C44.5 Skin of trunk | Basal cell carcinoma, NOS | 43.8 | UKBB |
| 7 | | C43.5 Malignant melanoma of trunk | . | 55 | UKBB |
| 7 | | C44.4 Skin of scalp and neck | . | 59 | UKBB |
| 7 | | C44.5 Skin of trunk, C44.6 Skin of upper limb, including shoulder | . | 54 | UKBB |
| 7 | | C97 Malignant neoplasms of independent (primary) multiple sites | . | 66 | UKBB |
| 8 | p.Glu76Asp | C71.2 Temporal lobe | Glioblastoma, NOS | 63.4 | UKBB |
| 9 | p.Gln79Arg | C50.9 Breast | . | 62 | UKBB |
| 9 | | 1869 Malignant neoplasm of testis, other and unspecified | . | 27.8 | UKBB |
| 10 | p.Glu258Asp | C20 Malignant neoplasm of rectum | Adenocarcinoma, NOS | 69.9 | UKBB |
| 11 | p.Leu262Phe | C20 Malignant neoplasm of rectum | . | 61.1 | MyCode |
| 12 | p.Arg265Gln | C19 Malignant neoplasm of rectosigmoid junction | Adenocarcinoma, NOS | 59.6 | UKBB |
| 12 | | C20 Malignant neoplasm of rectum | | 60 | UKBB |
| 12 | | C78.0 Secondary malignant neoplasm of lung; C78.7 Secondary malignant neoplasm of liver | | 60 | UKBB |
| 13 | p.Arg265Gln | C34.1 Upper lobe, bronchus or lung | Adenocarcinoma, NOS | 46.5 | UKBB |
| 13 | | C77.1 Intrathoracic lymph nodes | | 47 | UKBB |
| 14 | p.Arg265Gln | C61 Malignant neoplasm of prostate | Adenocarcinoma, NOS | 68.6 | UKBB |
| 15 | p.Ile282Val | C44.9 Malignant neoplasm of skin, unspecified | Basal cell carcinoma, NOS | 64.7 | UKBB |
| 16 | p.Val428Met | C74.1 Medulla of adrenal gland | Neoplasm, malignant | 41.2 | UKBB |
| 17 | p.Thr468Met | C44.5 Skin of trunk | Malignant melanoma, NOS | 41 | MyCode |
| 17 | | C44.3 Skin of other and unspecified parts of face | Lentigo maligna | 55 | MyCode |
| 17 | | C44.6 Skin of upper limb and shoulder; C44.7 Skin of lower limb and hip | Melanoma in situ | 59 | MyCode |
| 17 | | C44.6 Skin of upper limb and shoulder | Lentigo maligna | 44 | MyCode |
| 18 | p.Arg498Trp | C43.6 Malignant melanoma of upper limb, including shoulder | . | 69 | UKBB |
| 19 | p.Arg501Lys | C62.9 Testis, unspecified | Seminoma, NOS | 57.8 | UKBB |
| 20 | p.Met508Val | C44.3 Basal cell carcinoma ok skin of nose, C44.9 Basal cell carcinoma of skin, unspecified | . | 68 | BioMe |
| 21 | p.Met508Val | C18.1 Malignant neoplasm of appendix | . | 76.9 | MyCode |
| 22 | p.Gln510Arg | C44.3 Skin of other and unspecified parts of face | Basal cell carcinoma, NOS | 58.1 | UKBB |
| 22 | | C17.9 Small intestine, unspecified | Carcinoid tumour, malignant | 74.2 | UKBB |
| 22 | | C44.6 Skin of upper limb, including shoulder | Squamous cell carcinoma, NOS | 77.4 | UKBB |
| 22 | | C43.5 Malignant neoplasm of trunk | . | 73 | UKBB |
| 22 | | C44.4 Skin of scale and neck, C44.6 Skin of upper limb, including shoulder | . | 78 | UKBB |
| 22 | | C44.5 Skin of trunk | . | 74 | UKBB |
| 22 | | C75.9 Endocrine gland, unspecified | . | 78 | UKBB |
| 22 | | C78.6 Secondary malignant neoplasm of retroperitoneum and peritoneum, C78.7 Secondary malignant neoplasm of liver | . | 75 | UKBB |
| 22 | | C97 Malignant neoplasms of independent (primary) multiple sites | . | 78 | UKBB |
| 23 | p.Gln510Arg | C43.4 Malignant melanoma of scalp and neck | Lentigo maligna melanoma | 57.1 | UKBB |

**J.**

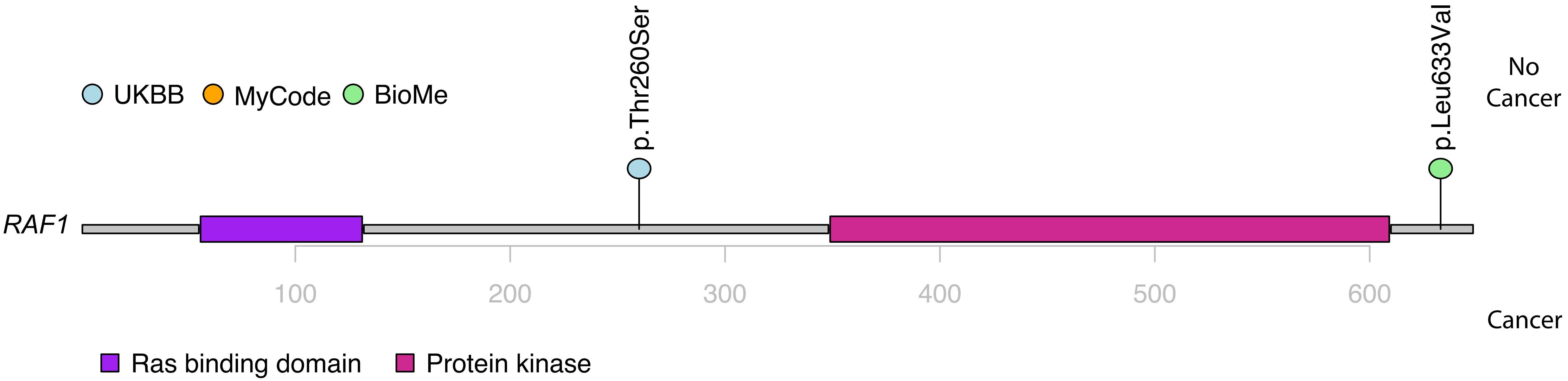

UKBB
MyCode
BioMe
p.Thr260Ser
p.Leu633Val
No Cancer
RAF1
100
200
300
400
500
600
Cancer
Ras binding domain
Protein kinase

**K.**

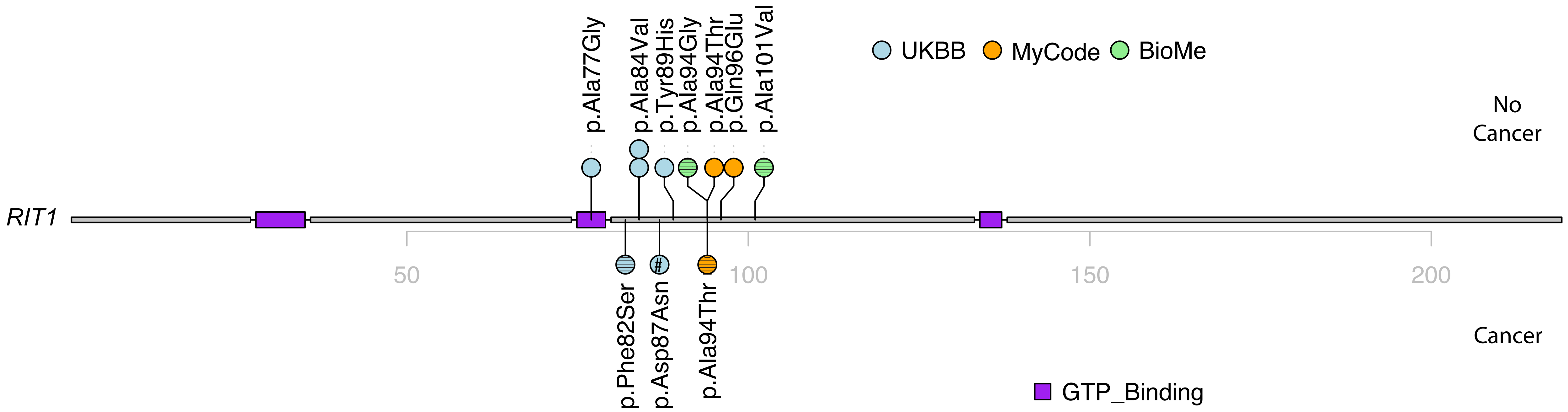


| | HGVS p. | Cancer | Histology | Age at diagnosis | Database |
|---|---|---|---|---|---|
| 1 | p.Phe82Ser | C44.9 Malignant neoplasm of skin, unspecified | Squamous cell carcinoma, NOS | 79.2 | UKBB |
| 1 | | C44.9 Malignant neoplasm of skin, unspecified | Basal cell carcinoma, NOS | 80.5 | UKBB |
| 2 | p.Asp87Asn | C34.1 Upper lobe, bronchus or lung; C34.9 Bronchus or lung, unspecified | Small cell carcinoma, NOS | 69.2 | UKBB |
| 2 | | C78.7 Secondary malignant neoplasm of liver | . | 69 | UKBB |
| 2 | | C79.3 Secondary malignant neoplasm of brain and cerebral meninges; C79.5 Secondary malignant neoplasm of bone and bone marrow | . | 70 | UKBB |
| 3 | p.Ala94Thr | C44.5 Unspecified malignant neoplasm of skin of other part of trunk | . | 72.6 | MyCode |
| 3 | | C44.2 Basal cell carcinoma of skin of unspecified ear and external auricular canal | . | 75.8 | MyCode |
| 3 | | C44.3 Squamous cell carcinoma of skin of other parts of face | . | 80.2 | MyCode |

L.

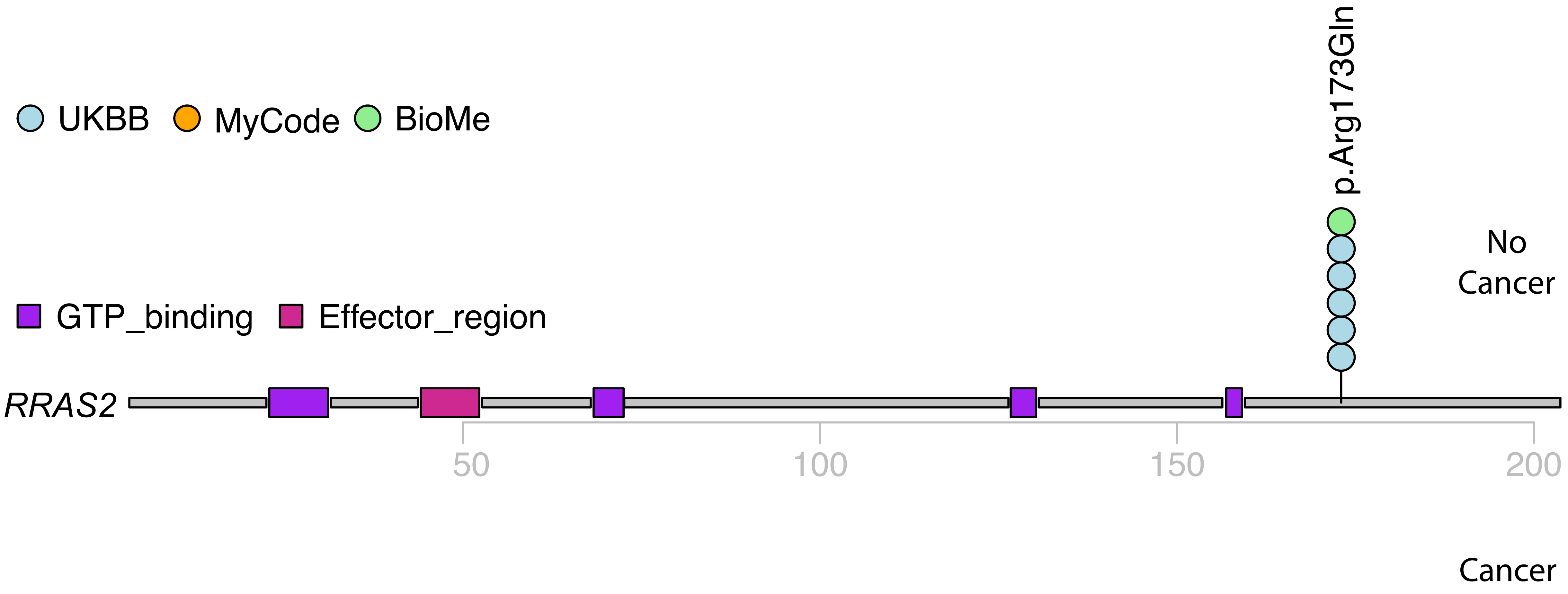

UKBB
MyCode
BioMe
p.Arg173Gln
No Cancer
GTP_binding
Effector_region
RRAS2
50
100
150
200
Cancer

**M.**

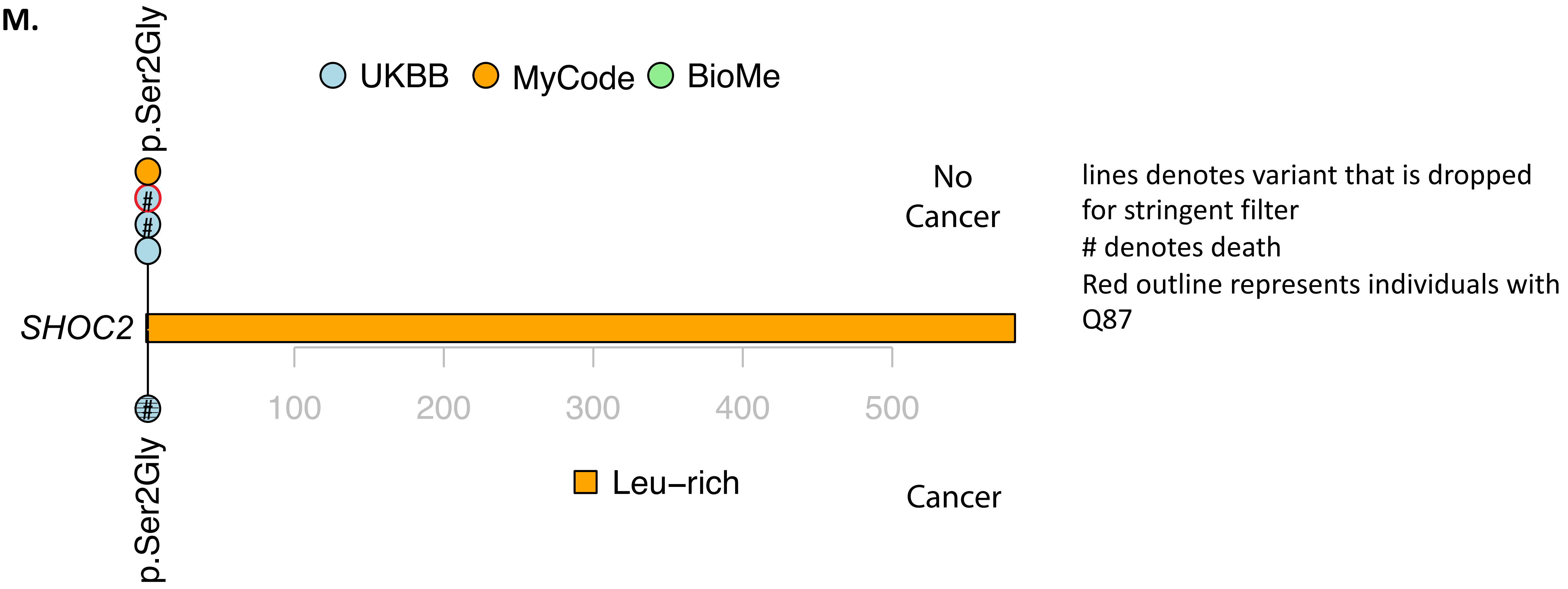


| | HGVS p. | Cancer | Histology | Age at diagnosis | Database |
|---|---|---|---|---|---|
| 1 | p.Ser2Gly | C34.2 Middle lobe, bronchus or lung; C34.9 Bronchus or lung, unspecified | Adenocarcinoma, NOS | 76.1 | UKBB |
| 1 | | C79.5 Secondary malignant neoplasm of bone and bone marrow | . | 77 | UKBB |

N.

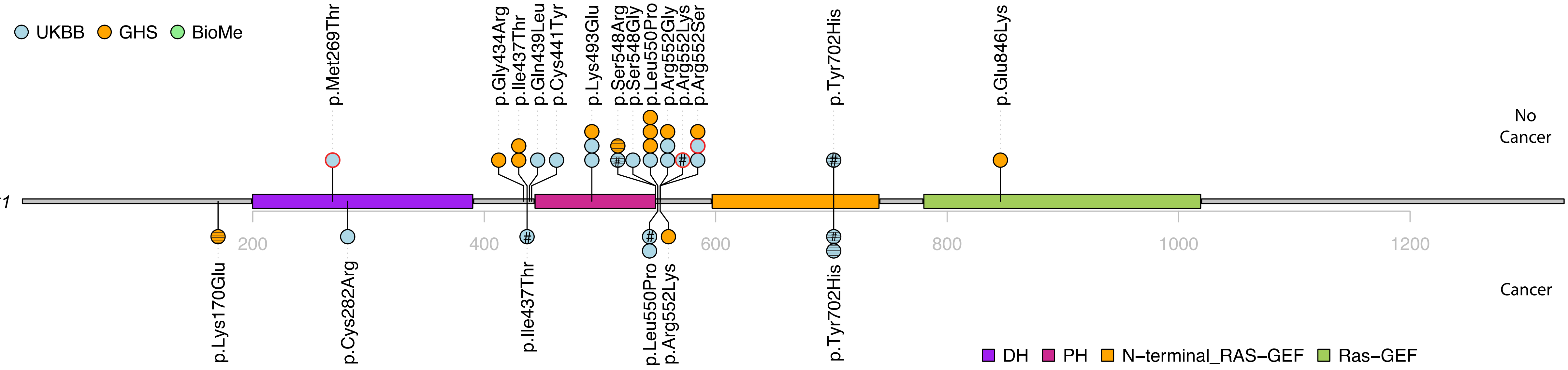


| | HGVS p. | Cancer | Histology | Age at diagnosis | Database |
|---|---|---|---|---|---|
| 1 | p.Lys170Glu | C56.9 Malignant neoplasm of unspecified ovary | Low-grade serous carcinoma | 67 | MyCode |
| 2 | p.Cys282Arg | C50.4 Upper-outer quadrant of breast | Infiltrating duct carcinoma, NOS | 50.8 | UKBB |
| 3 | p.Ile437Thr | C34.1 Upper lobe, bronchus or lung; C34.9 Bronchus or lung, unspecified | Adenocarcinoma, NOS | 68.7 | UKBB |
| 3 | | C79.3 Secondary malignant neoplasm of brain and cerebral meninges | . | 68 | UKBB |
| 3 | | C79.5 Secondary malignant neoplasm of bone and bone marrow | . | 69 | UKBB |
| 3 | | C80.9 Malignant neoplasm, unspecified | . | 68 | UKBB |
| 4 | p.Leu550Pro | C80 Malignant neoplasm without specification of site | . | 66.8 | UKBB |
| 4 | | C56 Malignant neoplasm of ovary | . | 67 | UKBB |
| 4 | | C78.6 Secondary malignant neoplasm of retroperitoneum and peritoneum; C78.7 Secondary malignant neoplasm of liver; C78.8 Secondary malignant neoplasm of other and unspecified digestive organs | . | 67 | UKBB |
| 4 | | C79.9 Secondary malignant neoplasm, unspecified site | . | 69 | UKBB |
| 5 | p.Leu550Pro | C44.1 Skin of eyelid, including canthus | Basal cell carcinoma, NOS | 67.3 | UKBB |
| 5 | | C44.3 Skin of other and unspecified parts of face | | 72 | UKBB |
| 6 | p.Arg552Lys | C44.9 Unspecified malignant neoplasm of skin, unspecified | . | 43.2 | MyCode |
| 7 | p.Tyr702His | C16.0 Cardia; C16.9 Stomach, unspecified | Adenocarcinoma, NOS | 65.6 | UKBB |
| 7 | | C15.0 Cervical part of oesophagus;C15.9 Oesophagus, unspecified | . | 66 | UKBB |
| 8 | p.Tyr702His | C44.2 Skin of ear and external auricular canal | Basal cell carcinoma, NOS | 72.1 | UKBB |

**O.**

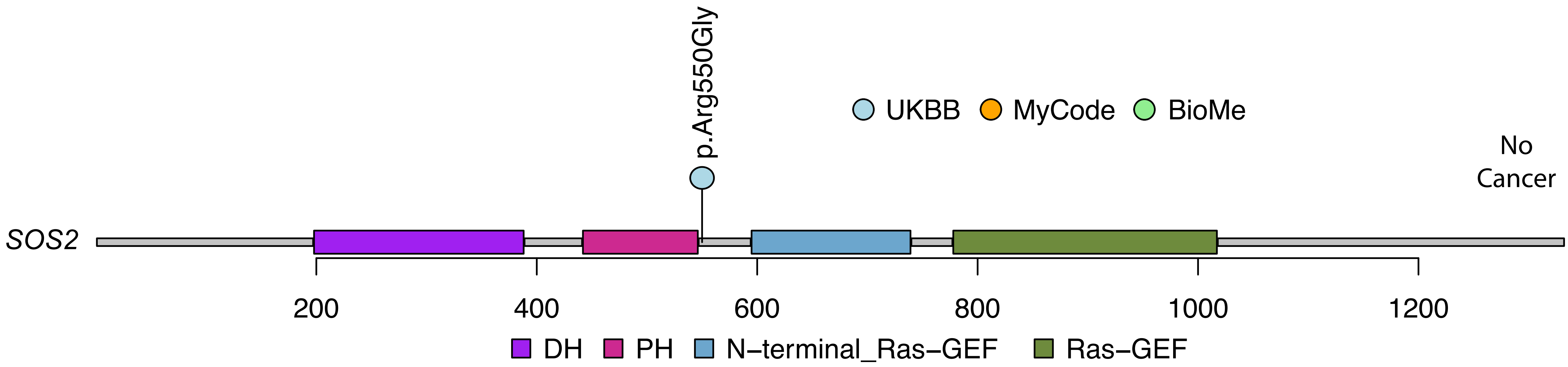
p.Arg550Gly
UKBB
MyCode
BioMe
No Cancer
SOS2
200
400
600
800
1000
1200
DH
PH
N–terminal_Ras–GEF
Ras–GEF

P.

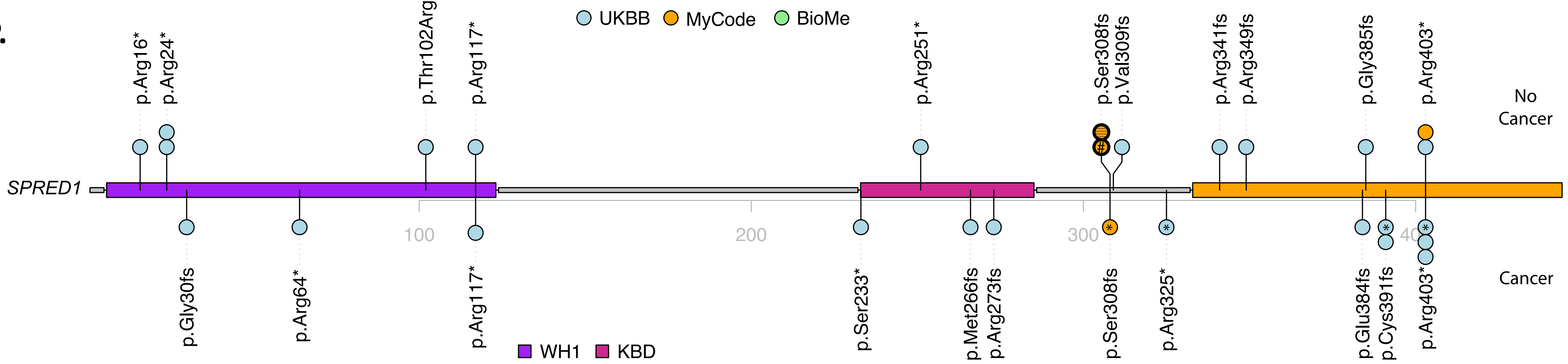


| | HGVS p. | Cancer | Histology | Age at diagnosis | Database |
|---|---|---|---|---|---|
| 1 | p.Gly30fs | C67.9 Bladder, unspecified; C67.4 Posterior wall of bladder | Papillary trans. cell carcinoma | 57.3 | UKBB |
| 2 | p.Arg64* | C17.0 Duodenum | Adenocarcinoma, NOS | 57.8 | UKBB |
| 2 | | C16.9 Stomach, unspecified | . | 58 | UKBB |
| 3 | p.Arg117* | C44.6 Skin of upper limb, including shoulder | . | 73 | UKBB |
| 4 | p.Ser233* | C44.9 Malignant neoplasm of skin, unspecified | Basal cell carcinoma, NOS | 67.1 | UKBB |
| 5 | p.Met266fs | C44.3 Skin of other and unspecified parts of face | Basal cell carcinoma, NOS | 44 | UKBB |
| 5 | | C61 Malignant neoplasm of prostate | Adenocarcinoma, NOS | 63.7 | UKBB |
| 6 | p.Arg273fs | C44.3 Skin of other and unspecified parts of face; C44.1 Skin of eyelid, including canthus | Squamous cell carcinoma, NOS | 69.6 | UKBB |
| 7 | p.Ser308fs | C44.0 Basal cell carcinoma of skin of lip | . | 56.4 | MyCode |
| 8 | p.Arg325* | C80 Malignant neoplasm without specification of site | Adenocarcinoma, metastatic, NOS | 56.7 | UKBB |
| 8 | | C16.9 Stomach, unspecified | . | 57 | UKBB |
| 8 | | C78.0 Secondary malignant neoplasm of lung; C78.2 Secondary malignant neoplasm of pleura; C78.6 Secondary malignant neoplasm of retroperitoneum and peritoneum;C79.5 Secondary malignant neoplasm of bone and bone marrow | . | 57 | UKBB |
| 9 | p.Glu384fs | C50.9 Breast, unspecified; C50.5 Lower-outer quadrant of breast | Infiltrating duct carcinoma, NOS | 58.6 | UKBB |
| 9 | | C80 Malignant neoplasm without specification of site | . | 64 | UKBB |
| 10 | p.Cys391fs | C34.9 Bronchus or lung, unspecified | Carcinoma, NOS | 73 | UKBB |
| 11 | p.Cys391fs | C43.4 Malignant melanoma of scalp and neck | Superficial spreading melanoma | 63.8 | UKBB |
| 12 | p.Arg403* | C50.8 Overlapping lesion of breast | Infiltrating duct carcinoma, NOS | 63.1 | UKBB |
| 12 | | C50.9 Breast, unspecified | . | 51 | UKBB |
| 13 | p.Arg403* | C64 Malignant neoplasm of kidney, except renal pelvis | Clear cell adenocarcinoma, NOS | 73.4 | UKBB |
| 13 | | C78.7 Secondary malignant neoplasm of liver; C79.5 Secondary malignant neoplasm of bone and bone marrow | | 73 | UKBB |
| 14 | p.Arg403* | C44.1 Skin of eyelid, including canthus | Basal cell carcinoma, NOS | 40.5 | UKBB |
